# The occurrence of cross-host species soil-transmitted helminth infections in humans and domestic/livestock animals: a systematic review

**DOI:** 10.1101/2025.04.26.25326220

**Authors:** Uniqueky Gratis Mawrie, Riviarynthia Kharkongor, María Martínez Valladares, Stella Kepha, Sitara S. R. Ajjampur, Rajiv Sarkar, Rachel Pullan

## Abstract

Zoonotic soil-transmitted helminths (STH), including *Ancylostoma ceylanicum*, *Ancylostoma caninum*, *Ancylostoma braziliense*, *Trichuris vulpis*, *Trichuris suis*, and *Ascaris suum*, are increasingly recognised as potential sources of human infection. Additionally, animals can act as carriers or reservoirs for human STH species. However, the extent of cross-host infection remains poorly understood, primarily due to reliance on morphological diagnostics. This review compiles data on the occurrence of cross-host STH infections, highlighting zoonotic STH in humans and human STH species in domestic and livestock animals. Following PRISMA guidelines, PubMed, Medline, and Web of Science were systematically searched from inception to December 2024. Inclusion criteria encompassed studies on cross-host STH infections confirmed by molecular methods. Exclusion criteria included experimental infection studies, studies involving wildlife, and those that did not find cross-host infection. Two independent reviewers assessed bias using AXIS and Joanna Briggs Institute appraisal tools. The protocol is registered with PROSPERO (CRD42024519067). The review screened 4197 titles and abstracts; and included 51 studies. *Ancylostoma ceylanicum* was the commonest zoonotic STH reported, predominantly in Southeast Asia. Human STH species (*Ancylostoma duodenale, Necator americanus, Trichuris trichiura* and *Ascaris lumbricoides*) were found in dogs, cats, and pigs. Studies examining both humans and animals together in shared environments showed STH presence in both populations. Case studies revealed gastrointestinal and dermatological effects in humans particularly infected with zoonotic hookworms. This systematic review highlights STH cross-host species infections underscoring the need for further One health epidemiological investigations of humans and domestic/livestock animals in sympatric environments to better understand the burden and explore the transmission dynamics of cross-host STH infections.

## Background

The soil-transmitted helminths (STH) in humans are neglected tropical diseases caused commonly by *Ascaris lumbricoides*, *Trichuris trichiura* and hookworms (*Ancylostoma duodenale, Necator americanus*). The World Health Organisation has specified aims to eliminate STH as a public health problem in 96% of endemic countries by 2030, targeting a reduction in heavy-to-moderate intensity infection prevalence in children to below 2% by preventive chemotherapy [1]. While widespread deworming initiatives have successfully reduced morbidity in most settings [2], persistent environmental contamination may contribute to continued reinfection, raising questions around sustainability [3,4]. In order to address environmental contamination, focus has been primarily directed towards improvements in water, sanitation, and hygiene (WASH) infrastructure and behaviour change communication to prevent human faecal contamination [5,6]. However, evidence suggest WASH’s effectiveness is low-to-moderate [7–9] and it may not effectively address all sources of environmental contamination.

Zoonotic helminths are globally prevalent in animals, but the frequency of their occurrence in humans and its public health implications remain largely unexplored. Most notably, dogs and cats serve as hosts for the hookworm species *Ancylostoma ceylanicum*, *Ancylostoma caninum*, *Ancylostoma braziliense*, in addition to *Trichuris vulpis*, while pigs host *Ascaris suum* and *Trichuris suis* - all of whom have zoonotic potential. Limited evidence of their presence in humans is due to the inability of morphological egg identification (until recently, the primary diagnostic tool) to differentiate between species. Nevertheless, genetic analyses suggests potential cross-host species infection of *Ascaris* and *Trichuris* between humans and pigs [10–13], including hybridisation between *Ascaris suum* and *Ascaris lumbricoides* [14,15]. Furthermore, it is also possible that animals can act either as carriers/transport hosts [16,17] or reservoirs of human STH species, potentially influencing the transmission dynamics.

Given that dogs’ and cats’ faeces commonly contaminate soil in areas where they roam freely [18–21], and that pigs are reared closely with humans in many low-and-middle-income settings, it is important to ascertain their role in maintaining STH transmission in human populations. Considering the potential challenges to disease control, posed by animal reservoirs, this study systematically reviews evidence of cross-host species infections of zoonotic STH (*Ancylostoma ceylanicum*, *Ancylostoma caninum*, *Ancylostoma braziliense*, *Ascaris suum*, T*richuris vulpis* and *Trichuris suis*) in humans and human STH (*Ancylostoma duodenale*, *Necator americanus, Trichuris trichiura* and *Ascaris lumbricoides*) in domestic/livestock animals. It aims to understand the geographical distribution and the extent of cross-host species infections to explore the role of animals in the transmission of STH in endemic settings.

## Methods

### Search strategy and selection criteria

Following the Preferred Reporting Items for Systematic Review and Meta-Analyses (PRISMA) guidelines [22] (S1 Checklist), a systematic review on cross-host species infections of STH between humans and animals (domestic/livestock) was conducted. The protocol of this systematic review was registered with PROSPERO (registration ID CRD42024519067). The review was carried out by two independent reviewers, UM and RK. UM formulated the research questions, developed inclusion and exclusion criteria, and build the search strategy. Both reviewers independently handled study selection, data extraction, quality and bias assessments, resolving differences through discussions with RS. UM took the lead in the interpretation and reporting the findings.

### Database search

Three databases-PubMed, Medline, and Web of Science, were searched for studies published since inception to December 2024. The search strategy was designed to include evidence on “Cross-host species infection” between humans and animals defined as the occurrence of zoonotic STH species, namely *Ancylostoma ceylanicum*, *Ancylostoma caninum*, *Ancylostoma braziliense*, *Ascaris suum*, *Trichuris vulpis* and *Trichuris suis* in humans. It also included occurrences of human STH species that is, *Ancylostoma duodenale*, *Necator americanus*, *Trichuris trichiura* or *Ascaris lumbricoides* in domestic/livestock animals. BOOLEAN operators ‘OR’ and ‘AND’ were used to refine the search strategy. The list of search terms and details of the search strategy applied to extract studies from each database is described in S1 Table.

### Study selection

Original studies employing molecular methods to confirm STH species in cross-host species infections were considered for inclusion. Since this review aims to establish the occurrence (not necessarily the prevalence) of cross-host species infections, both community-based studies and studies conducted in hospital settings were included. We included only papers that explicitly reported cross-host species infections, excluding those that investigated but did not find evidence of such infections. Experimental studies (artificially-induced infections), studies reporting helminths in wildlife animals, studies that did not use molecular methods were excluded. Other forms of publications such as editorials, or review papers were also excluded but bibliography was checked for further references. Unpublished and grey literature were not assessed. *Strongyloides stercoralis*, although classified as an STH, was excluded from this review due to its unique features: under-reporting due to auto-infection [23], diagnostic limitations related to Kato-katz method [24], and Ivermectin as the preferred treatment and preventive deworming [25]; hence was excluded from this review.

### Data extraction

The studies identified from all the three databases were exported to Zotero (Zotero 6.0.36) [26] for management and referencing. Rayyan software [27] was used for study organisation and duplicate detection, with manual screening. After removing duplicates, titles and abstracts were screened. Studies with unclear eligibility were included for further assessment. Full-texts of eligible studies were retrieved. Information on the year of publication, species detected, animal hosts, country, diagnostic method used, sample size, number of samples tested by morphological examination, number of samples tested by molecular method, number of samples tested as positive, prevalence (with 95% confidence intervals, when applicable), clinical symptoms and case description (for case studies) were extracted (S2 Table). If participants had travelled to an endemic region, then the place of origin and the place travelled were also recorded (S3 Table). No pooled analysis was conducted.

### Quality assessment

Risk of bias was assessed using the Appraisal tool for Cross-sectional studies (AXIS) [28] for community-based studies and the Joanna Briggs Institute (JBI) Critical Appraisal tools for cross-sectional studies and case reports [29] for hospital-based studies. AXIS consists of 20 items evaluating study design, reporting quality, and bias risk. The JBI tool for cross-sectional studies assesses sample criteria, subject descriptions, measurement validity, confounding factors identification, and statistical analysis appropriateness. For case reports, the JBI tool evaluates clarity of case descriptions, suitability of diagnostic methods, consideration of confounding factors, and reliability of conclusions. All eligible studies were included regardless of the quality score.

## Results

### Search results

The process of selecting eligible studies using the PRISMA guidelines is presented in Fig 1.

**Fig 1.**
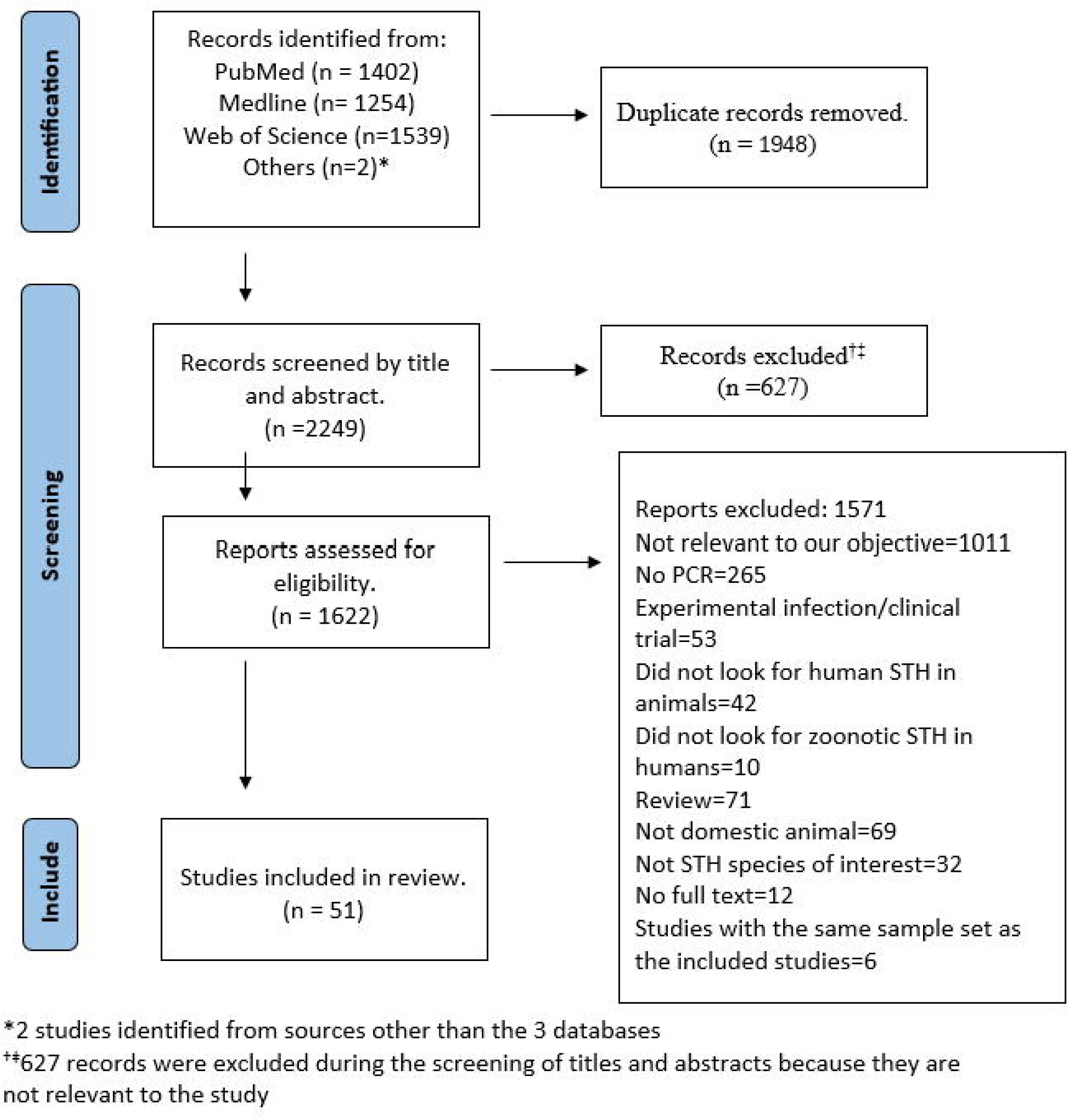
Flow diagram showing the screening and selection of studies using the Preferred Reporting Items for Systematic Reviews and Meta-Analyses (PRISMA).

The search identified 4197 studies. After removing 1948 duplicates, 2249 records were screened for titles and abstracts, of which 627 were excluded. A total of 1622 full-text articles were evaluated for eligibility against the pre-determined eligibility criteria. Fifty-one studies were included in this review, after excluding 1011 studies that were not relevant to the study objective, 265 studies that did not use molecular methods for identification of STH species, 53 experimental infection studies, 31 studies that only included unrelated helminth species, 69 studies involving only wild animals and 12 records for which full-text was not available (Fig 1). The 12 records for which the full texts were unavailable were published between 1949 and 2001. Given the age of these publications and the likelihood that they did not employ molecular diagnostic methods for species confirmation, efforts to contact the authors were not undertaken. The detailed reasons for excluding the papers after full-text assessment is explained in the S2 Table.

### Study characteristics

Thirty-five (68·6%) of the 51 studies that met the eligibility criteria were community-based while 16 (31·4%) studies were conducted in hospital settings. Majority of the studies (96·1%, 49 of 51) were published in the last decade (2010-2022) (S1 Fig). Studies conducted across 28 countries revealed the presence of cross-host species STH infections in humans and animals. Predominantly, 62·7% (32 of 51) of these studies were from 12 countries in Southeast Asia (SEA), with a smaller proportion from South America (11·8%, 6 of 51), Europe (9.8%, 5 of 51), Africa (7.8%, 4 of 51), South Asia (7.8%, 4 of 51) and Oceania (3·9%, 2 of 51) (Fig 2-3 and S2-S3 Figs).

**Fig 2.**
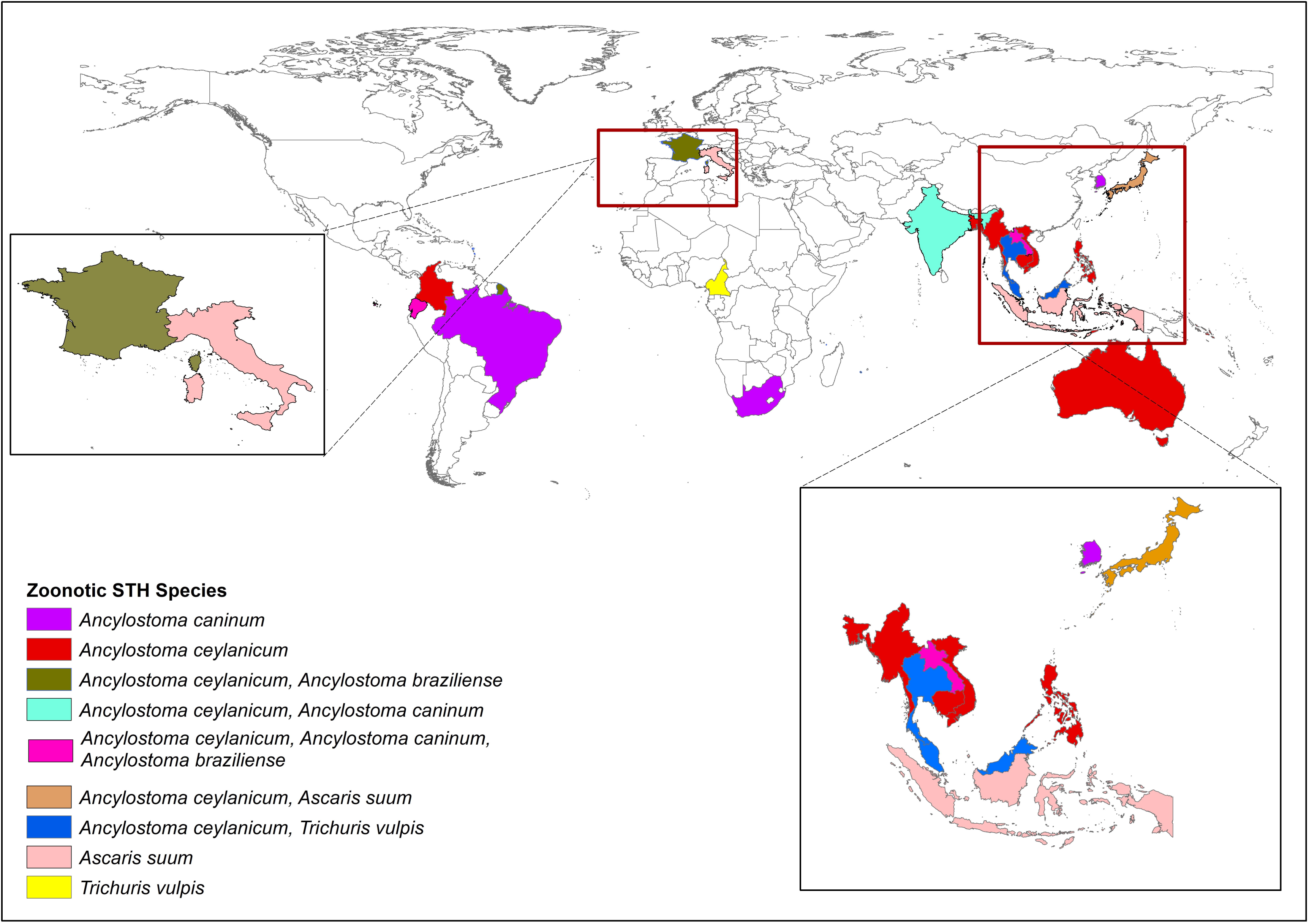
Distribution of studies reporting the occurrence of zoonotic STH across Southeast Asia, South Asia, Oceania, Africa, Europe and South America.

**Fig 3.**
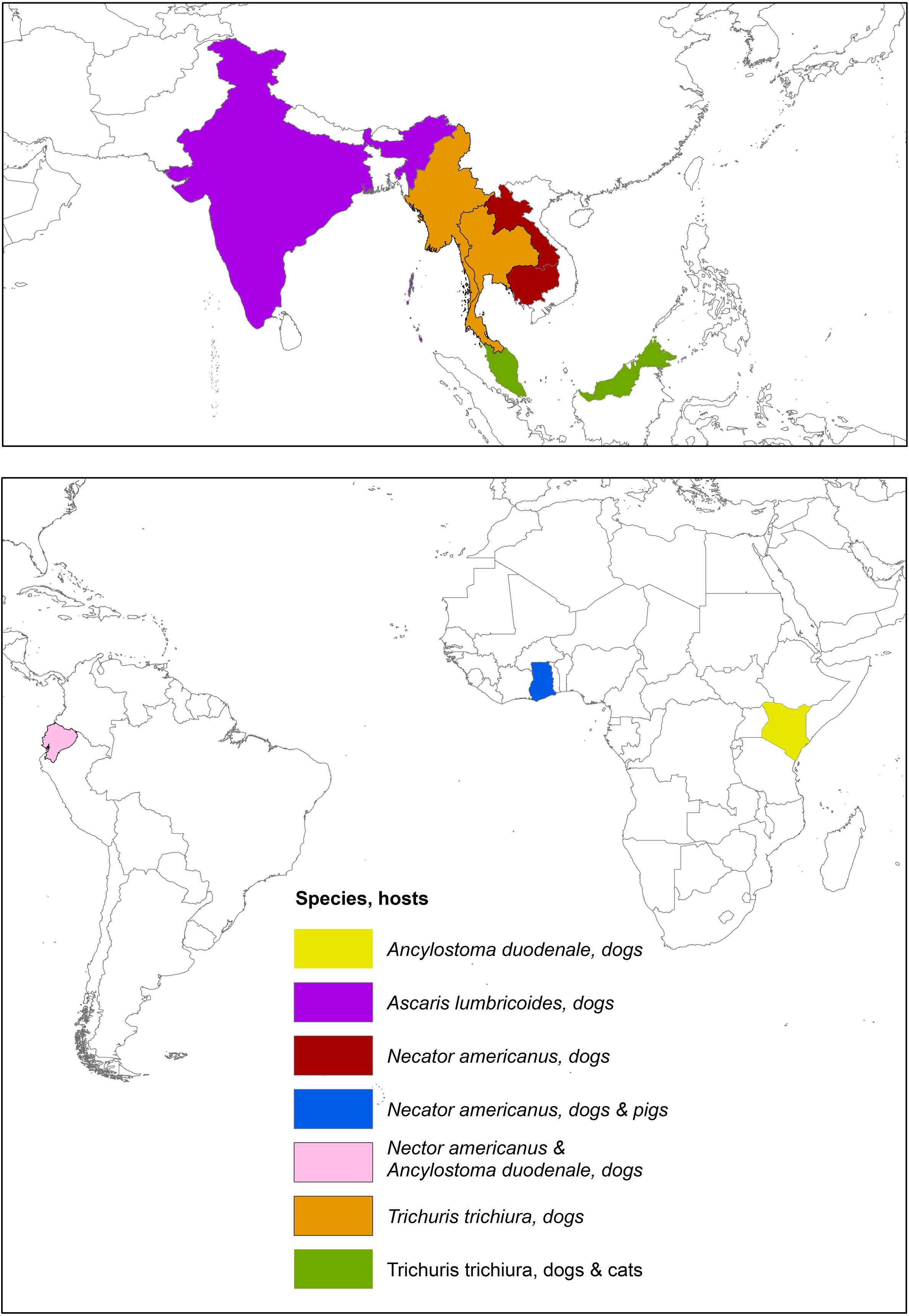
Distribution of studies reporting the occurrence of human STH species in animal hosts.

Among the community-based studies, the majority (75·0%, 27 of 36) utilised cross-sectional designs. A smaller percentage were integrated into larger programs (5·5%, 2 of 36) [30,31], while others employed alternative designs, including cluster-randomised trial (2.8%, 1 of 36) [32], multicentre drug efficacy trial (2.8%, 1 of 36) [33], testing/validation of diagnostic tools or treatment (5.6%, 2 of 36) [34,35], cohort study design (2.8%, 1 of 36) [36] or retrospective analysis of archived samples from previous studies (2.8%, 1 of 36) [37]. Rural populations were the focus of the majority of community-based studies (63.9%, 23 of 36) [37–45], with some studies (11.1%, 4 of 36) specifically targeting tribal or indigenous communities [37,39,41,42]. Additionally, 22·2% (8 of 36) focused on children, particularly preschool and school-aged groups, with a subset conducted in daycare centres and schools (5·6%, 2 of 36) [34,46,47]. Other specific study locations included communities around temples [48], refugee camps [30,31], and tea-growing communities [49]. Site selection methods varied, including accessibility-based approaches [39,41,43,50] and consideration of contact with domestic animals [43,49]. Sampling methodologies encompassed simple random sampling of households [50,51], random selection of villages/schools [41,52], random sampling of participants [53], purposive [45] and convenience sampling [54] (Table 1).

**Table 1.**
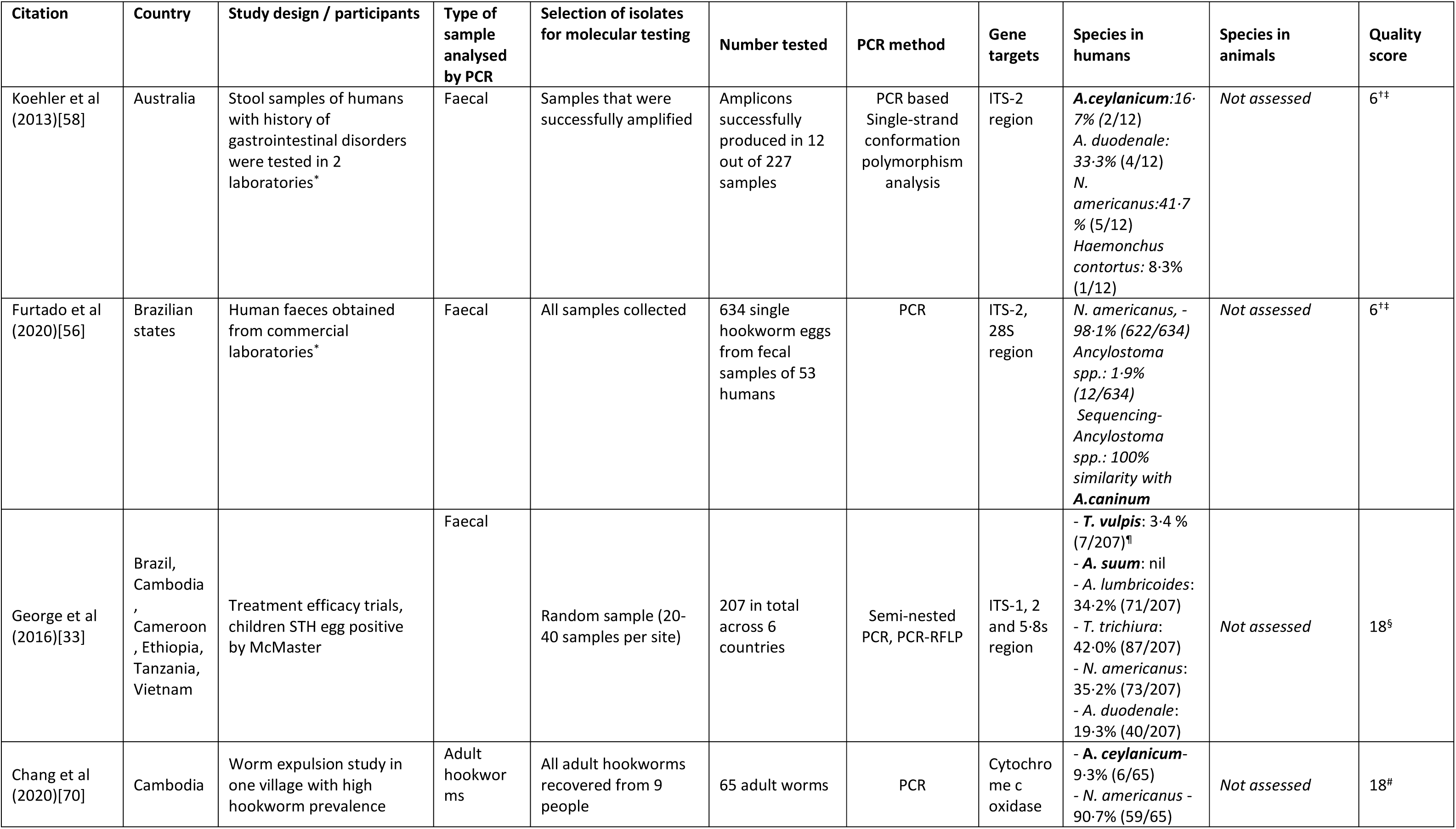
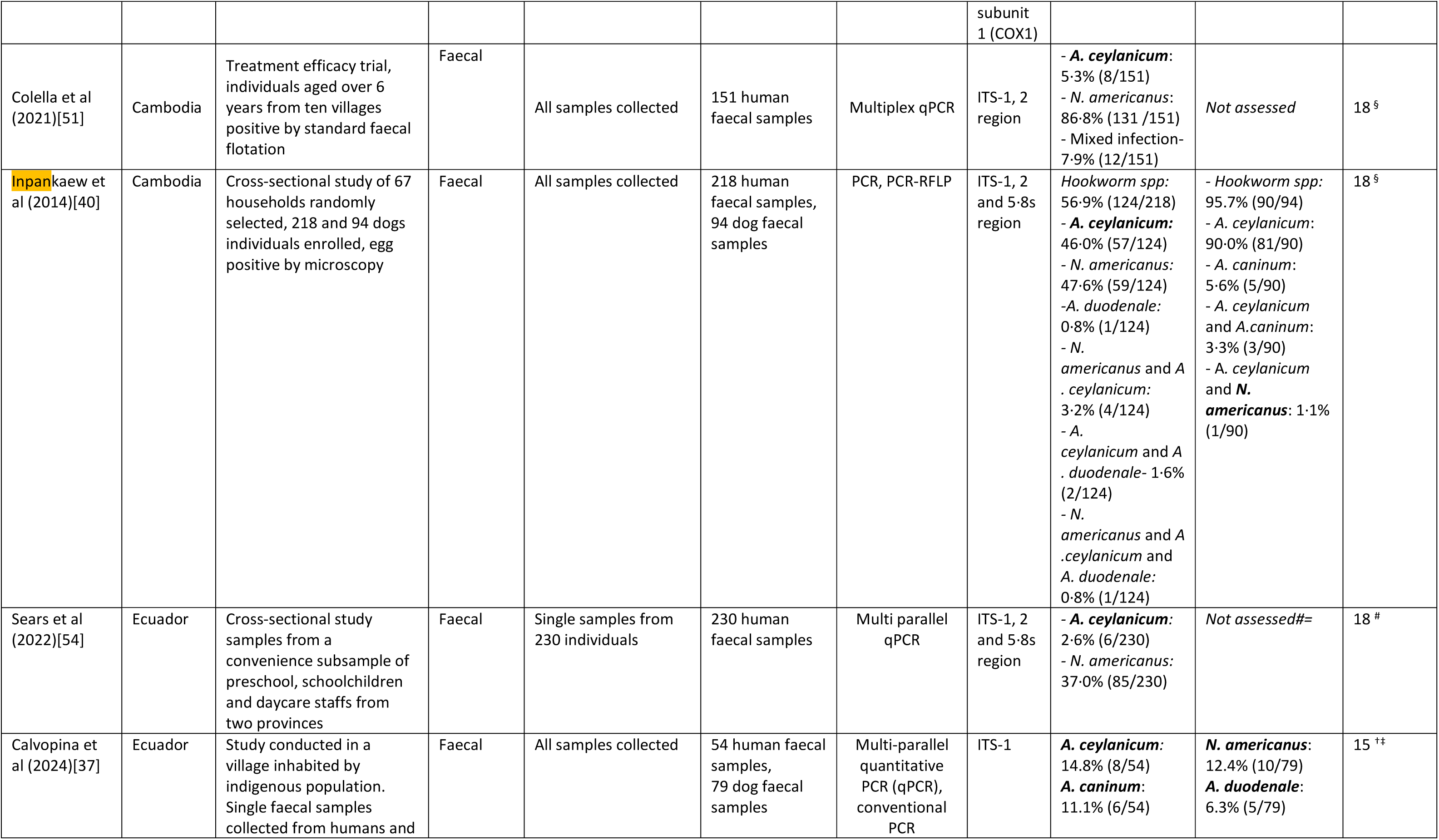
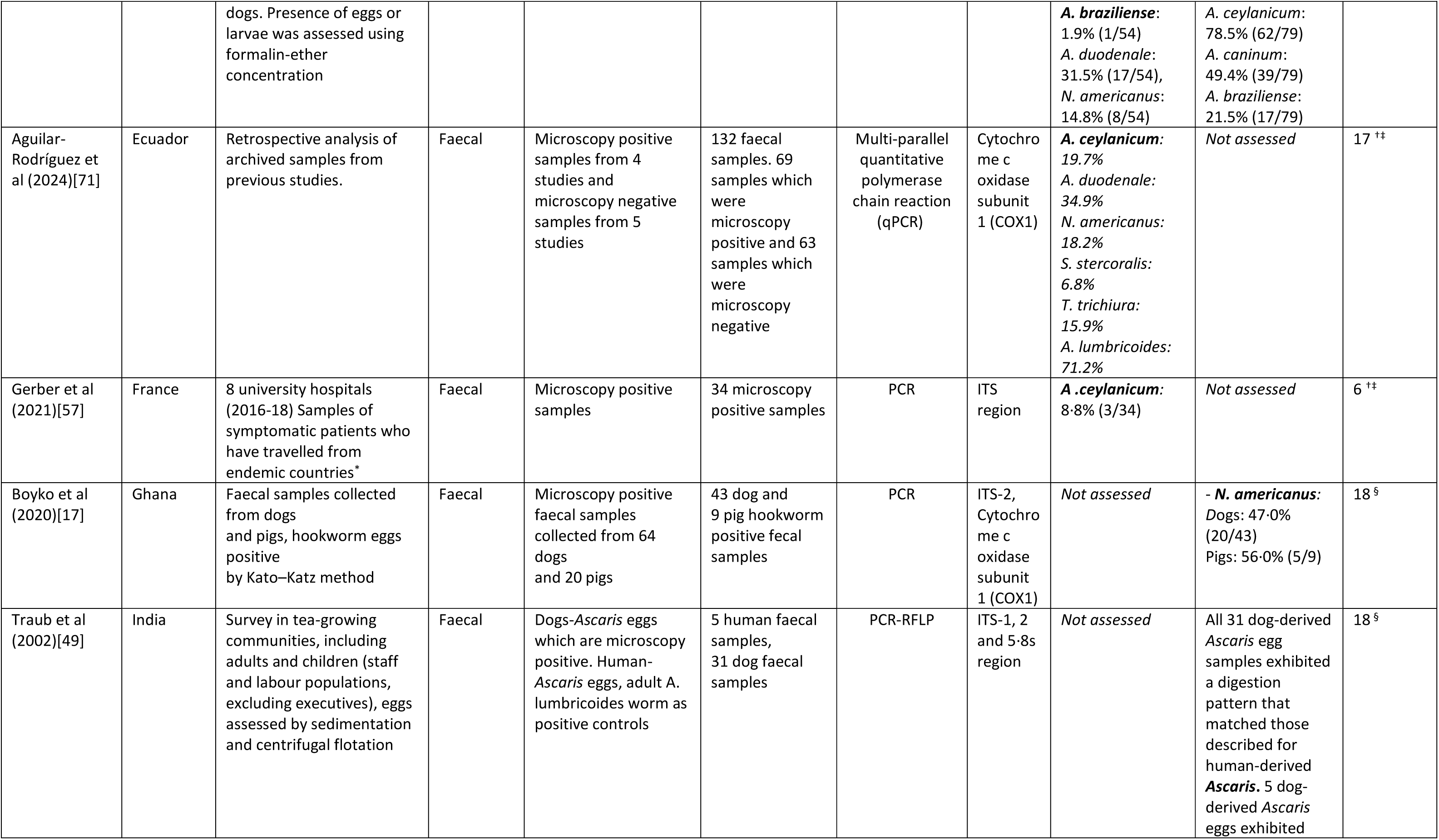
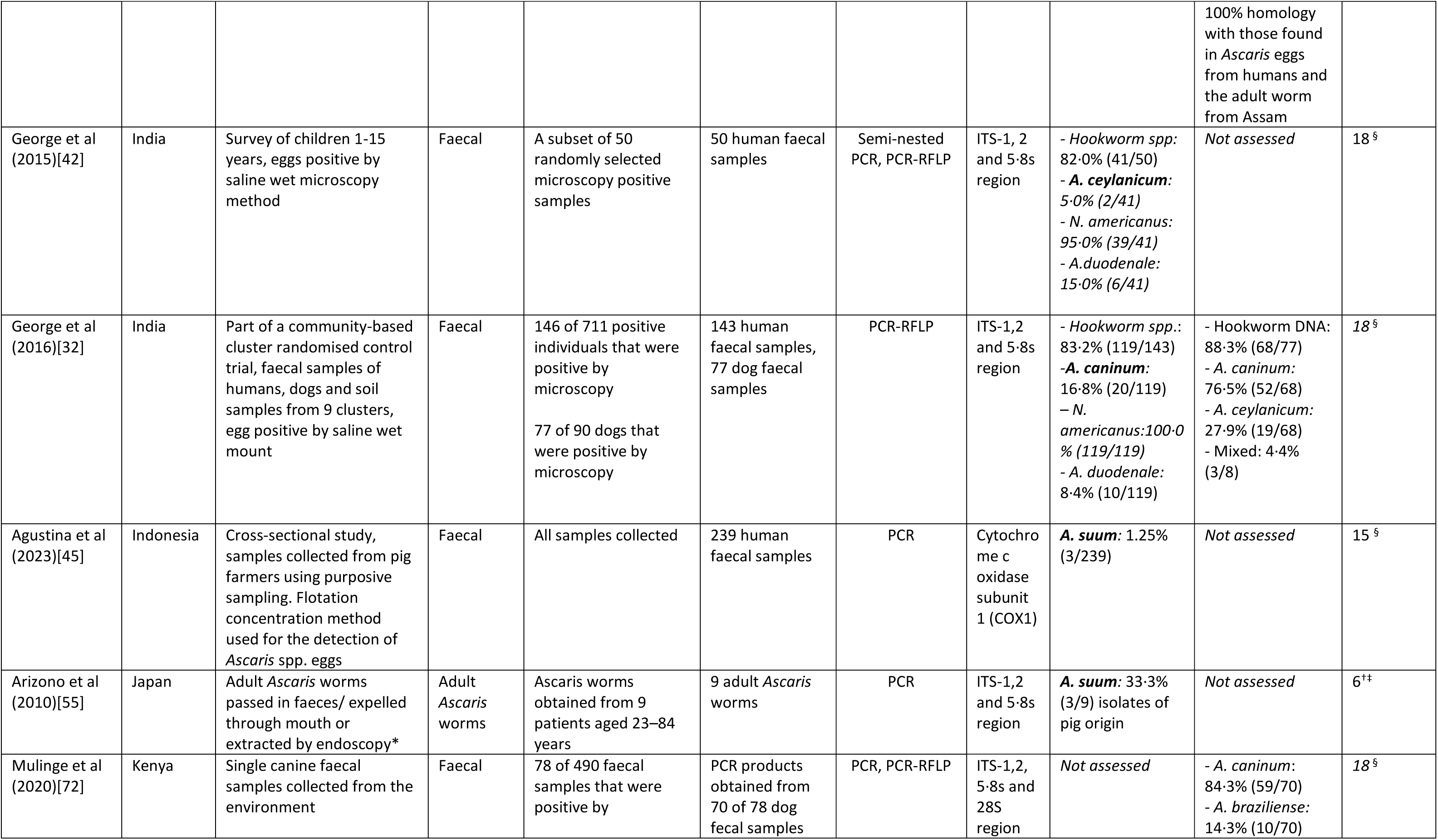
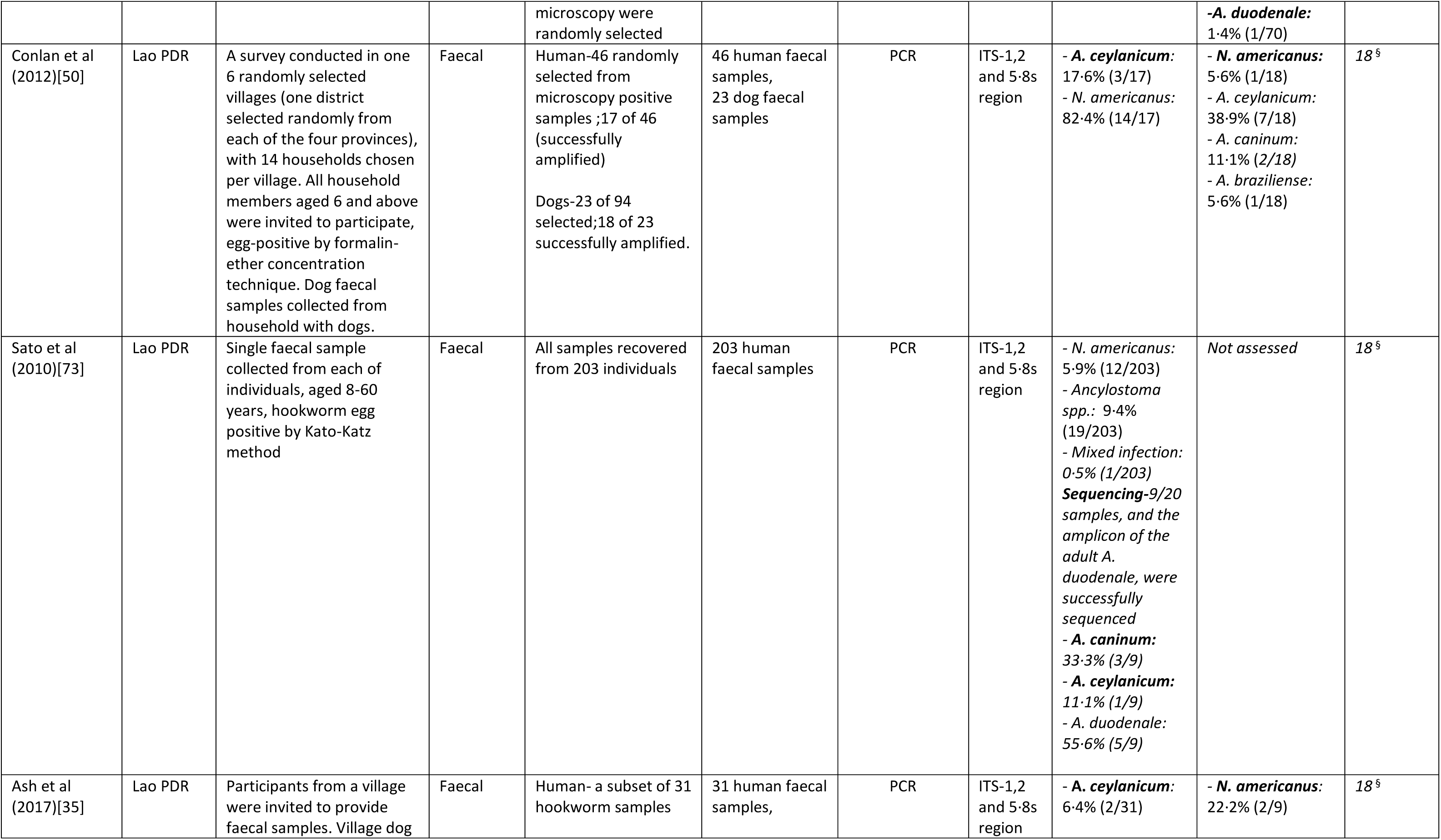
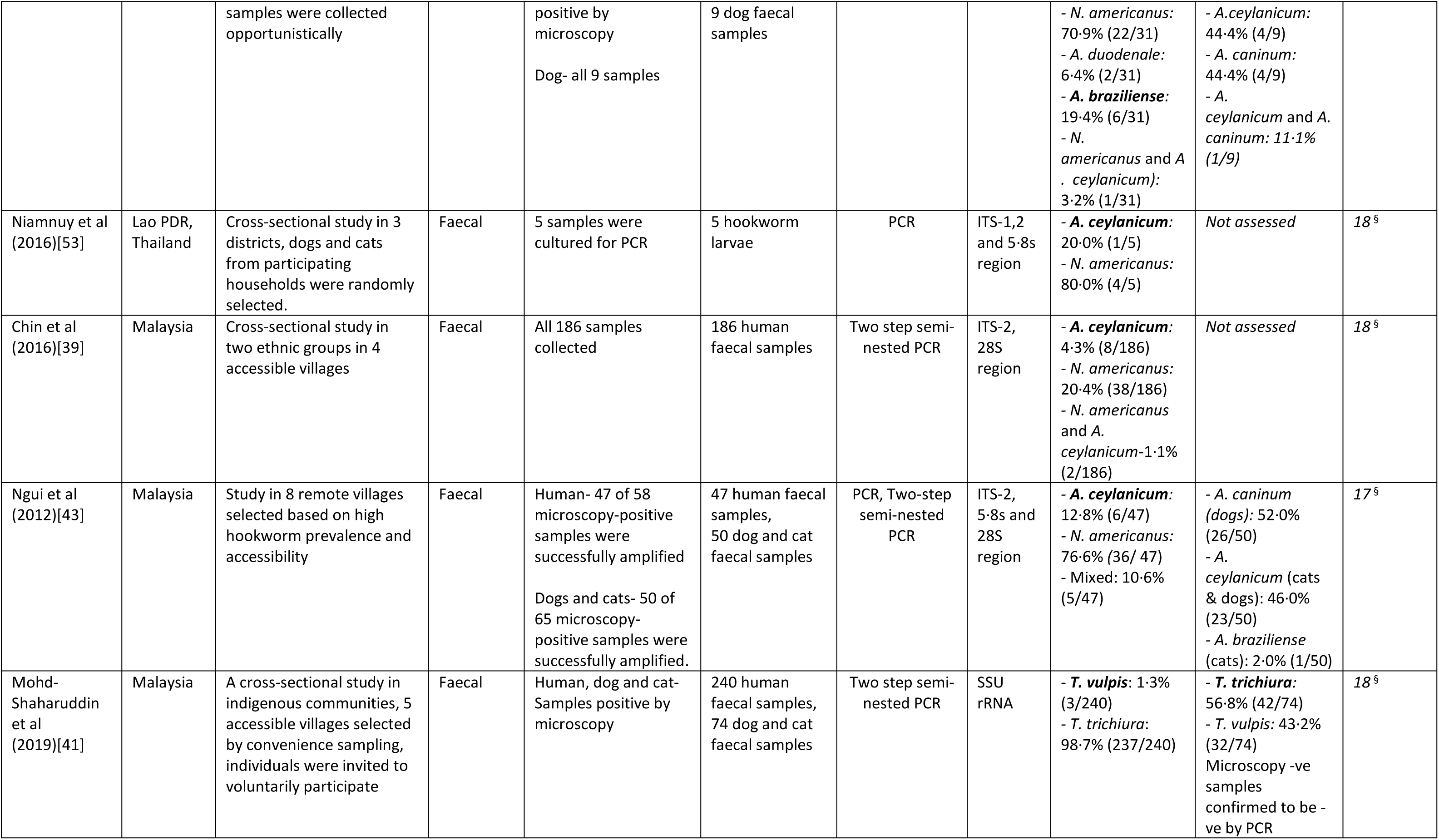
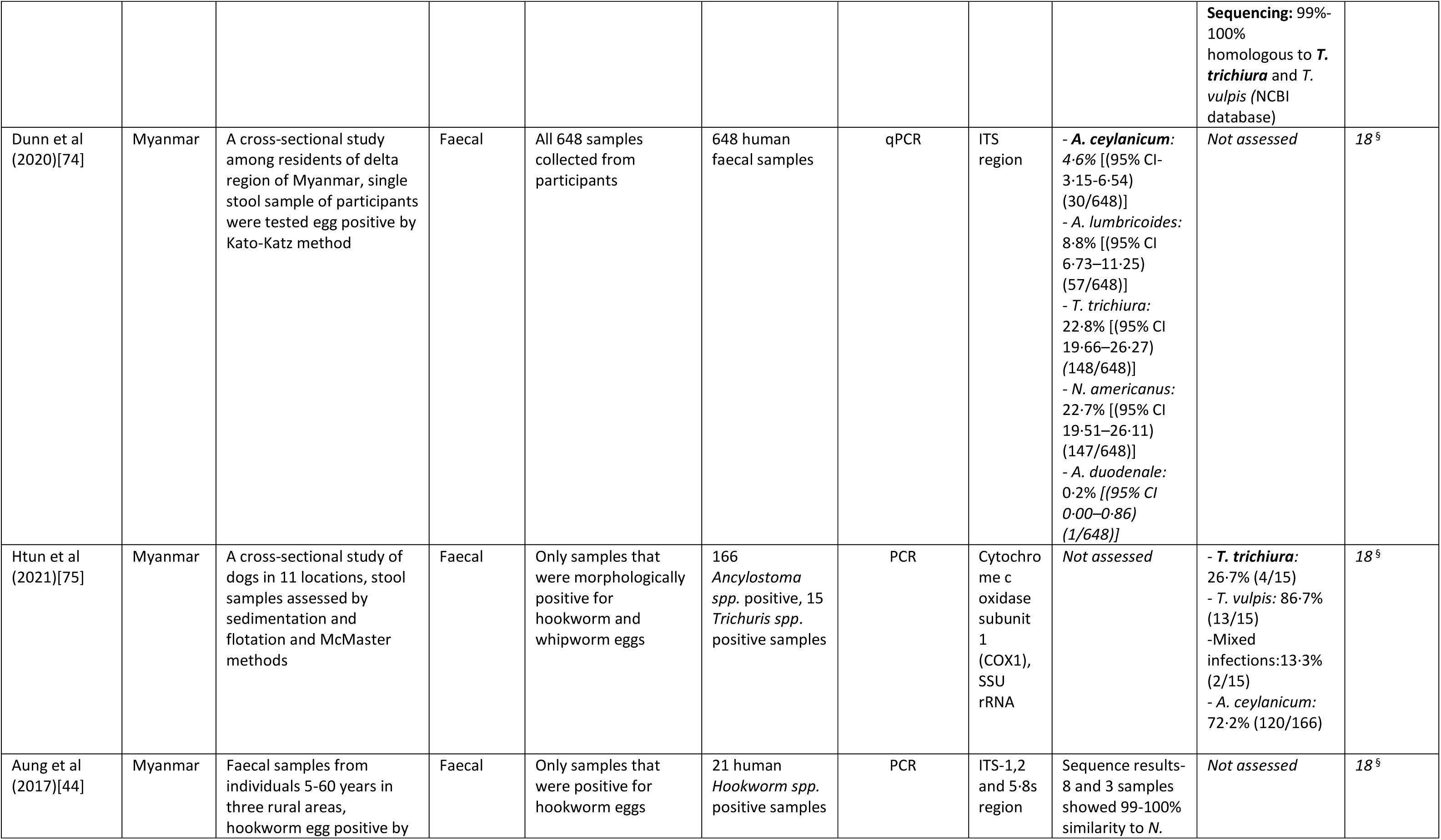
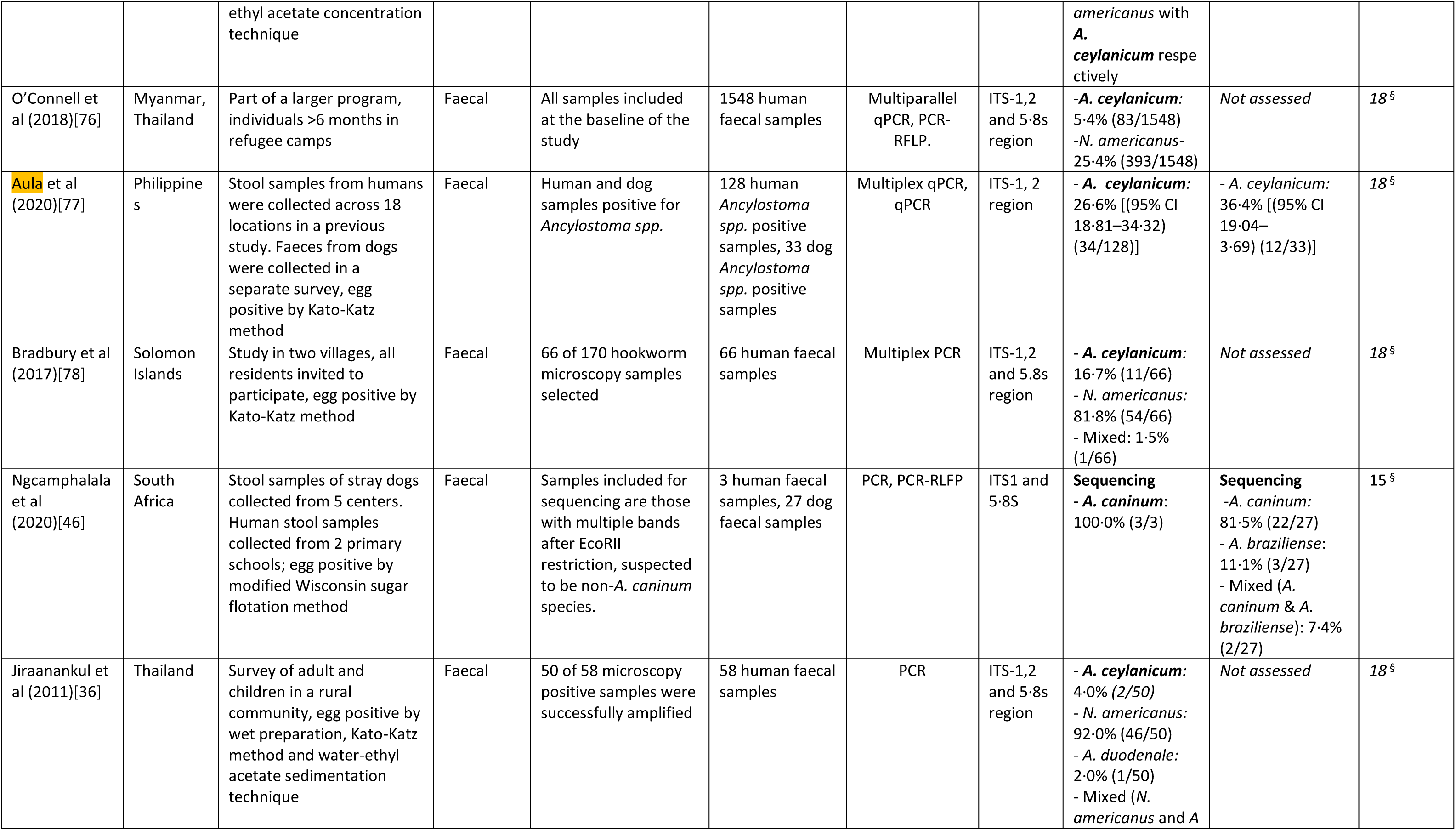
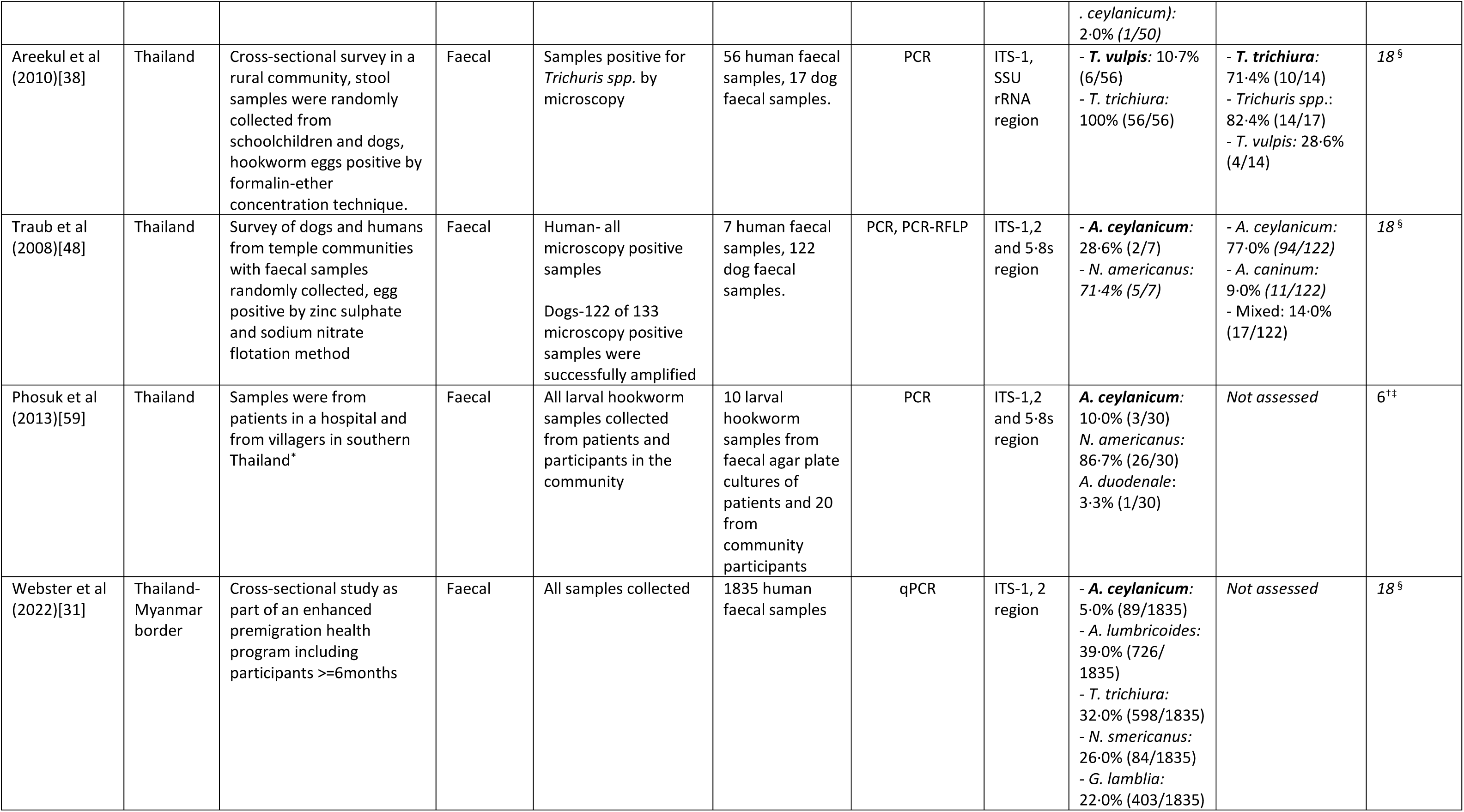
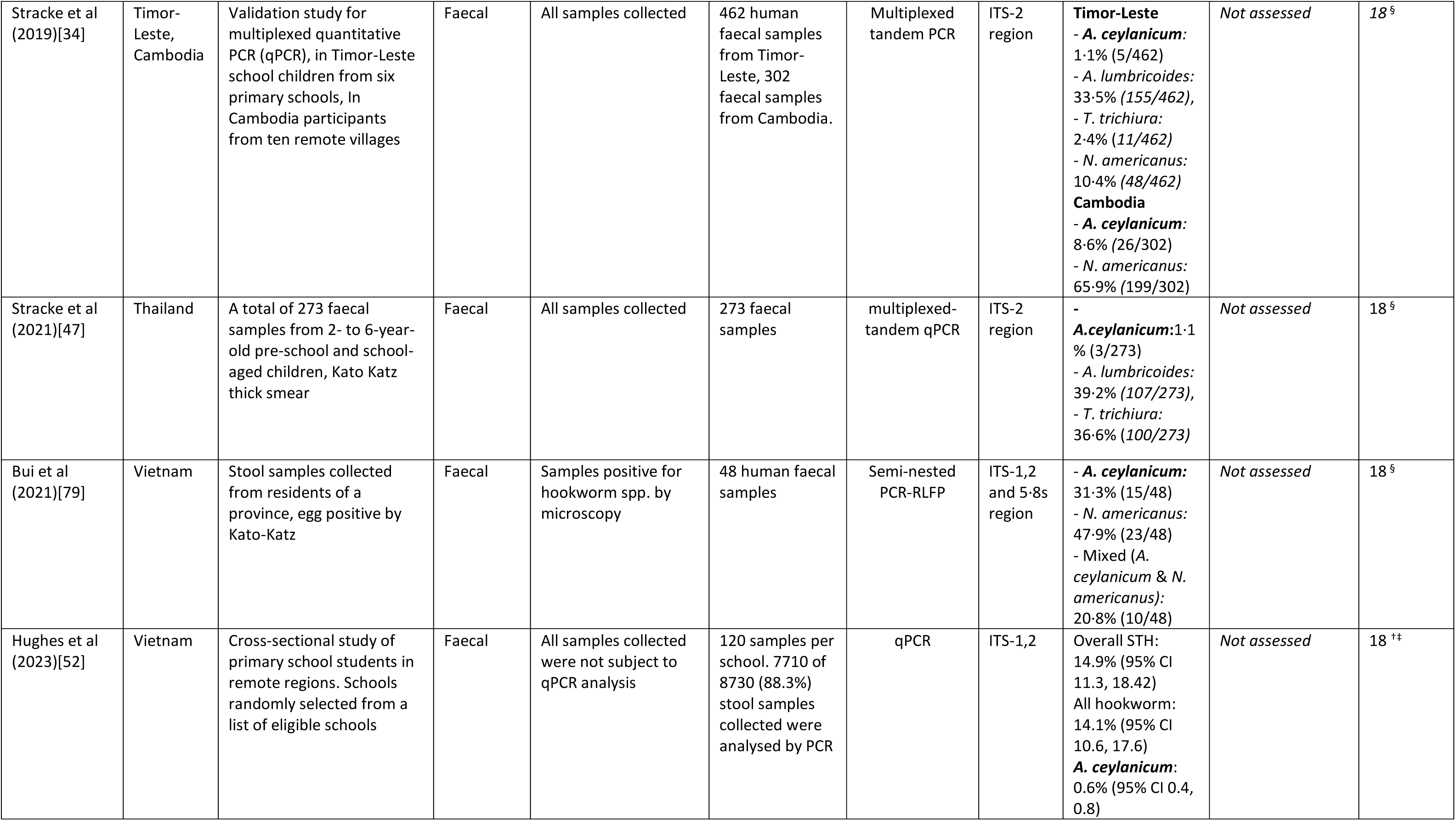
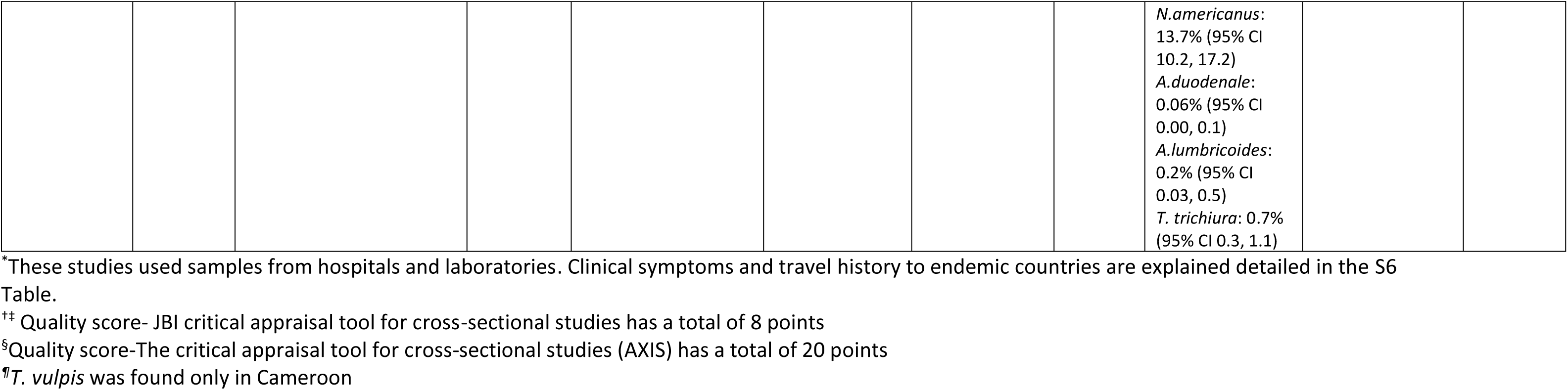
Characteristics of eligible studies reporting zoonotic STH species in humans and/or human STH in animals.

Among hospital-based studies, 73·3% (11 of 15) followed a case-study design, while five studies (33·3%, 5 of 15) utilised preexisting laboratory or clinical faecal samples [55–59], with one study sampled from both a hospital and a village [59]. These studies included symptomatic patients, with samples comprising of adult worms [60–65] and faeces [64,66–69]. Five studies (three from France and two from Japan) reported travel histories to endemic countries such as Thailand [64], Lao PDR [64,67], Myanmar [68], Malaysia [67], India [67], West Indies [66], Pakistan, Cote d‘Ivoire, Colombia, Pakistan, French Guiana [57] and Papua New Guinea [67]; while three mentioned contact with animals [60,62,63] (Table 2 and S6 Table).

**Table 2.** Summary of case studies and hospital-based studies included in the review.

| Citation | Country of origin | Clinical history/symptoms | Method of diagnosis | Type of sample analysed by PCR | Number of cases | Species detected | Travel history/animal contact | Quality score* |
| --- | --- | --- | --- | --- | --- | --- | --- | --- |
| Nath et al (2024)[69] | Bangladesh | Recurrent diarrhea and weakness | PCR | Faecal | One case | *A. ceylanicum* | - | 5 |
| Poppert et al (2017)[80] | Colombia | Loss of vision | PCR, sequencing | Adult worm was destroyed during surgical removal.<br>intraoperative<br>rinsing fluid was<br>used | One case | *A. ceylanicum* | Not recent travel recorded | 8 |
| Brunet et al<br>(2015)[68] | France | Fever, vomiting,<br>dyspnoea, bloody<br>diarrhoea and weight loss.<br>Pruritic erythematous<br>macules on buttocks while<br>in Myanmar | PCR | Faecal | One case | *A. ceylanicum* | Returned from<br>Myanmar | 7 |
| Joncour et al<br>(2012)[66] | France | Itchy rash, persistent<br>pruritis | PCR, DNA<br>sequencing | Larvae from skin<br>scrapings | One case | *A. braziliense* | Returned from the<br>West Indies | 7 |
| Romano et al<br>(2021)[62] | Italy | Abdominal pain, vomiting,<br>bloating | PCR | Adult worm | One case | *A. suum* | History of rearing<br>chickens and pigs | 7 |
| Dutto M et al<br>(2013)[63] | Italy | Found one worm in his<br>stool | PCR-RFLP | Adult worm | One case | a hybrid genotype-*A.*<br>*suum/lumbricoides* | No history of travel,<br>pig farmer | 7 |
| Yoshikawa et al<br>(2018)[67] | Japan | Three cases had<br>abdominal pain, diarrhoea | PCR | Faecal | 4 cases | *A. ceylanicum* | Case 1-returned<br>from Malaysia<br>Case 2-returned<br>from Papua New<br>Guinea<br>Case 3-returned<br>from Lao PDR<br>Case 4-returned<br>from India | 7 |
| Nishioka et al<br>(2024)[65] | Japan | Asymptomatic | PCR-RFLP | Worm collected<br>by colonoscopy | One case | *A. suum* | - | 7 |
| Jung et al<br>(2020)[60] | South<br>Korea | Moderate eosinophilia | PCR | Worm | One case | *A. caninum* | Patient owns a dog | 6 |
| Kaya et al<br>(2016)[64] | Japan | Intermittent diarrhoea,<br>eosinophilia. | PCR | Adult worm | One case | *A. ceylanicum* | Returned from<br>Thailand and Lao<br>PDR | 7 |
| Ngui et al<br>(2014)[61] | Malaysia | Upper GI bleed (blood in<br>stool) | PCR, DNA<br>sequencing | Worm | One case | *A. ceylanicum* | - | 5 |

\* Quality score- JBI critical appraisal tool for case studies has a total of 8 point

Polymerase chain reaction (PCR) methods commonly targeting genes such as Internal transcribed spacer-1,2 regions (ITS), mitochondrial cytochrome c oxidase subunit I (COX1), 5·8S, 18S and 28S ribosomal ribonucleic acid (rRNA) were used for species confirmation. In community studies, 44·4% (16 of 36) limited PCR testing to microscopy-positive samples, while 30·6% studies (11 of 36) analysed all samples (Table 1). In some cases (8·3%, 3 of 36), a subset of microscopy-positive samples was randomly selected for PCR [34,42,50]. PCR was conducted in faeces (80·4%, 41 of 51 studies) and in adult worms (15·7%, 8 of 51 studies). The largest surveys were in Myanmar and Thailand, focussing on refugees and employing PCR on all samples [30,31].

### Quality assessment of studies

Majority (88·9%, 32 of 36) of the community-based studies scored 17 points or higher out of a total of 20 points, indicating high quality. Among hospital-based studies, 87·5% (14 of 16) scored 6 points or higher out of a total of 8 points, also reflecting good quality. Details of the quality assessment of the studies are in S4, S6, S7 Tables.

### Evidence of cross-host species infections

#### Occurrence of zoonotic STH in humans

Notably, *Ancylostoma ceylanicum* was the most-reported zoonotic STH species, appearing in 66·7% (34 of 51) of the studies. Its distribution spanning 16 countries, mainly in SEA (61·8%, 21 of 34), with additional occurrences noted in the Solomon Islands, India, Bangladesh, Ecuador, Colombia, France and Australia (Fig 2). Case studies were reported from Japan, France, Malaysia, Bangladesh and Colombia [61,64,67–69,80] (Table 2).

Twenty studies reported its frequency as a percentage of hookworm positives with proportions ranging from 2·6% [54] to 46·0% [40]. Although *Ancylostoma ceylanicum* generally appeared to be a minor infection, in four studies its positivity rate was comparable to *Necator americanus* [37,40,71,79]. Ten studies, which analysed all collected samples by PCR, found its contribution to the overall prevalence, ranging from 1·1% to 14·8% [30,31,34,37,39,51,54,59,70,81] (Table 1). Other zoonotic STH species observed in SEA included *Ancylostoma braziliense* (3·4%, 1 of 29 studies), *Trichuris vulpis* (6.9%, 2 of 29 studies), *Ancylostoma caninum* (3.4%, 1 of 29 studies), and *Ascaris suum* (6.9%, 2 of 29 studies), although their presence was more sporadic.

Reports of zoonotic STH in humans were found to be documented infrequently across other regions globally. In Africa, *Ancylostoma caninum* [46] and *Trichuris vulpis* [33] in humans were identified in one study each. Likewise, South America witnessed four studies reporting *Ancylostoma ceylanicum* [37,54,71,80] and one reporting *Ancylostoma caninum* [56]. In Oceania, *Ancylostoma ceylanicum* was reported in two studies [58,78]. In South Asia, it was documented in two studies [42,69], and *Ancylostoma caninum* [32] was reported in another. Finally, in Europe, two studies identified *Ancylostoma ceylanicum* [57,68], two studies found *Ascaris suum* [62,63], and one study detected *Ancylostoma braziliense* [66] (Fig 2 and S2 Fig).

*Ancylostoma braziliense* was reported in a tribal population in Laos [35], with another occurrence reported in a case study in France [66]. *Ancylostoma caninum* was identified in human populations in India [32], Brazil [56], South Africa [46], Lao PDR [73] and in a case study in South Korea [60].

*Trichuris vulpis* was detected in human faeces in two studies in SEA and one in Cameroon [33,38,41]. *Ascaris suum* in humans occurred in one community study in Indonesia [45], in two case studies conducted in Italy, along with another study in a hospital in Japan where samples from patients were identified as *Ascaris suum* [55,62,63].

#### Morbidities linked with zoonotic STH infections

Clinical case studies documented zoonotic STH infections in humans, presenting with gastrointestinal disturbances (diarrhoea, vomiting, blood in stools, constipation), fever, eosinophilia, difficulty in breathing, and weight loss [57,58,61,62,64,67,68]. One case of *Ancylostoma braziliense*, confirmed by PCR on two larvae obtained from skin scrapings, presented with itchy rash and persistent pruritic but no gastrointestinal symptoms [66]. In another case of *Ancylostoma ceylanicum* identified by PCR of larvae, involved pruritic erythematous macules and gastrointestinal symptoms [68]. Two asymptomatic cases were detected through routine testing after returning from abroad [67] (Table 2 and S6 Table).

#### Occurrence of human STH species in animals

Only nine (17·6%) studies [35,37,38,40,41,50,59,72,75] have reported the presence of human STH species in animals. Instances where human STH species were identified in animals involved dogs, cats, and pigs. The occurrence of *Trichuris trichiura* (among *Trichuris spp.* infected dogs and cats) ranged from 26·7% to 71·4% in Malaysia, Thailand, and Myanmar [38,41,75].

*Necator americanus* was found in 12.4% (10/79 samples) of dog stools in Ecuador [37], 1 of 18 hookworm samples in Laos [50], and in 47% of dogs (20/43 samples) and 56% of pigs (5/9 samples) in Ghana [17]. *Ancylostoma duodenale* was found in 6.3% (5/79 samples) of dog stools in Ecuador [37] and 1 of 70 dog hookworm-positive dog samples in Kenya [72]. In India, 31 dog-derived *Ascaris* egg samples matched the digestion pattern of human-derived *Ascaris* by PCR-Restriction Fragment Length Polymorphism, with five showing 100% homology with human *Ascaris* eggs and the adult worm [49]. Human STH in animals were not reported from other parts of the world (Fig 3 and S3 Fig).

### Human-animal sympatric studies

Among the studies described above, eleven studies systematically explored human and animal populations in shared environments, predominantly investigating dogs and cats with humans [32,35,37,38,40,41,43,46,48–50]. When *Trichuris vulpis* was detected in humans, notably dogs and cats, in the same environment showed high infection rates ranging from 28·6% [38] to 43·2% [41]. Interestingly, in these studies, both dogs and humans were also found to be infected with *Trichuris trichiura* with prevalences ranging 56·8% −71·4% and 98·7-100·0% respectively [38,41]. Similarly, when humans were infected with *Ancylostoma ceylanicum,* [35,40,43,48,50] correspondingly, dogs showed high infection rates ranging from 38·9% to 90·0%. In India, *Ancylostoma caninum* was found in humans, with dogs also showing a high infection rate of 76·5% [32]. Furthermore, when dogs were infected with *Necator americanus* with prevalences 1·1% - 22·2%, humans also exhibited high infection rates ranging from 47·6% to 82·4% [35,40,50]. Similarly, in areas where dogs were infected with *Ancylostoma duodenale*, humans were also found to be infected with the same species [37].

## Discussion

This systematic review consolidates evidence of STH cross-host species infections, shedding light on their distribution and diversity. Analysing 51 studies on infections from stool and whole worm samples, the notable presence of zoonotic hookworm infections in humans is highlighted, with *Ancylostoma ceylanicum*, being a key source of human infections in SEA. Additionally, other zoonotic STH, such as *Ancylostoma caninum*, *Ancylostoma braziliense*, *Ascaris suum* and *Trichuris vulpis* were reported sporadically worldwide suggesting an under-recognised global issue. Despite genetic evidence of cross-host infections of *Ascaris* and *Trichuris spp.* between humans and pigs, fewer studies have explored this. The role of animals as reservoirs or carriers for human STH remains under-investigated. Our findings stress the importance of sampling in sympatric environments to better understand these dynamics and underscore the need for representative data, integrating molecular methods and fostering cross-sector collaboration to address animal reservoirs.

Majority of the human studies in this review focussed on zoonotic hookworms, particularly *Ancylostoma ceylanicum* predominantly reported in SEA, but also in other regions. *Ancylostoma caninum* and *Ancylostoma braziliense*, were less commonly reported, although they are known to be widely distributed among dogs in tropical regions [82–89]. With dog ownership averaging 130 dogs per 1000 people globally [90], current data may underestimate the true occurrence of these zoonotic hookworm infections in humans. This raises the possibility that observed human hookworm infections could include contributions from *Ancylostoma ceylanicum*, *Ancylostoma caninum* and *Ancylostoma braziliense*, misidentified as *Ancylostoma duodenale*. Although zoonotic hookworm infections in humans rarely constitute a major proportion of overall STH or hookworm positives, even low levels of cross-host species infection may have the potential to maintain transmission between humans and animal reservoirs, perpetuating the risk of re-infection and hindering efforts to achieve disease elimination. The emergence of reduced anthelmintic efficacy in humans [91], combined with resistance to benzimidazole [92,93] as seen in *Ancylostoma caninum* in dogs in the United States of America [93–95], highlights potential future challenges.

The review also sheds light on the zoonotic potential of *Trichuris vulpis* and *Ascaris suum* although in a limited number of studies. *Trichuris vulpis*, primarily found in dogs was detected in human faecal samples in Cameroon [33], Malaysia[41] and Thailand [38]. Similarly, *Ascaris suum*, a pig helminth, was identified in humans in Italy [62,63], Japan[55] and Indonesia [45]. Despite genetic studies demonstrating hybridisation between *Ascaris lumbricoides* and *Ascaris suum* [10,96,97] and between *Trichuris trichiura* and *Trichuris suis*, confirming distinct species with high genetic variation [98,99], suggesting cross-species transmission dynamics between humans and pigs [12,13,98,100], studies exploring these interactions remain scarce. Given the prevalence of small-scale pig farming globally, the role of pigs as potential reservoirs for zoonotic STH warrants further investigation. Additionally, coprophagy in animals can facilitate parasite transmission making them an important part of the transmission cycle.

Only a limited number of studies have investigated the presence of human STH in animals, but this lack of investigation does not imply their absence. Studies found the presence of *Trichuris trichiura* [38,41,75]*, Necator americanus* [17,35,37,40,50], *Ancylostoma duodenale* [37,72] and *Ascaris lumbricoides* in dogs, cats and pigs.

The presence of animal reservoirs could significantly hinder eliminate efforts, which largely rely on mass drug administration (MDA). This necessitates exploring STH transmission through a One Health lens. For example, a modelling study demonstrated that extending MDA to dogs could significantly reduce human *Ancylostoma ceylanicum* prevalence to less than 1·0% with just 25-50% deworming coverage of dogs by 2030 [101]. Additionally, the studies of humans and animals in shared environments also highlighted coinfection of zoonotic and human STH species in both populations, further emphasising the potential for cross-host transmission in areas of human-animal coexistence. Understanding the dynamics of cross-host STH infections remains complex. Evidence is needed to confirm whether zoonotic and human STH can complete their life cycles in alternative hosts. Zoonotic STH eggs may mature and reproduce in humans or pass through the human body without causing any harm, and human STH eggs may behave similarly in animals. Although uncertain whether other zoonotic STHs can complete their life cycles in humans, the presence of viable zoonotic hookworm eggs and adult worms seen in the studies of this review suggests potential for onward transmission. Additionally, animals may also serve as mechanical transmitters or transport hosts [16,17] likely contributing to STH transmission

Hospital based studies on symptomatic patients and those with travel history confirmed the clinical relevance of zoonotic STH, particularly *Ancylostoma ceylanicum*, *Ascaris suum*, *and Ancylostoma caninum*. Common clinical manifestations include gastrointestinal disturbances, fever, eosinophilia, respiratory difficulties, and weight loss [57,58,60–62,64,67,68]. Zoonotic hookworm infections are also linked to cutaneous larva migrans, though evidence is limited because of underreporting and infrequent investigation of zoonotic hookworms in most settings. Although these studies may not represent the broader distribution of infection, the presence of patent eggs and worms confirmed by molecular analyses highlights the potential of zoonotic STH to cause certain morbidities in humans.

This study has important limitations. Despite, recent community-based studies indicating increasing interest in exploring the occurrence of cross-host species infections, in this review we could not establish its true extent due to heterogeneity in study designs, diverse sampling strategies, limited geographic representativeness, and variations in sample selection criteria for molecular analyses. Sample bias, especially in studies with voluntary participation, hindered drawing comprehensive conclusions on the burden of cross-host species infections in humans and animals.

In conclusion, the scarcity of information regarding cross-host species infections at present could be attributed to limited exploration, related to morphological examination of eggs, hindering species identification. Studies focusing on *Ancylostoma ceylanicum* are concentrated in SEA, targeting rural communities with high hookworm prevalence and including households with domestic animals. To address this gap, it is essential to strengthen surveillance by incorporating molecular methods and fostering cross-sector collaboration. Expanding research to diverse geographical regions beyond SEA and conducting studies in sympatric environments, particularly in areas with high human-animal interaction, are crucial steps forward. As global initiatives aim to reduce STH morbidity by 2030, improved data on cross-host species infections are essential for informed interventions and improved public health outcomes.

## Data Availability

The data used in this analysis is provided in the supplementary materials.

## Supporting Information

**S1 Table: Detailed search strategy and records retrieved from each database.**

**S2 Table: Reasons for excluding studies after full-text assessment against eligibility criteria.**

**S3 Table: Description of variables extracted.**

**S4 Table: Quality assessment of studies that used samples from hospitals or laboratories using the AXIS critical appraisal tool for cross-sectional studies.**

**S5 Table: Quality assessment of 5 studies that used samples from hospitals or laboratories using the JBI checklist for cross-sectional studies.**

**S6 Table: Characteristics of eligible studies collected samples from hospitals/clinics/laboratories. S7 Table: Quality assessment of 9 case reports using the JBI checklist for case reports.**

**S1 Fig: Number of studies per year and the cumulative total of included studies.**

**S2 Fig: Number of studies by country reporting zoonotic STH in humans.**

**S3 Fig: Number of studies by country reporting human STH in animals**

