## Supplementary material for "The occurrence of cross-host species soil-transmitted helminth infections in humans and domestic/livestock animals: a systematic review": S1 Table. Detailed search strategy and records retrieved from each database..docx

| **Database searched** | **Search strategy** | **Records** |
| --- | --- | --- |
| Medline | ((("Ascaris lumbricoides" or "Trichuris trichiura" or "Ancyclostoma duodenale" or "Necator americanus") and ("small ruminants" or cow or cattle or pig or swine or dogs or canine or cats or feline)) or (("Ascaris suum" or "Ancylostoma ceylanicum" or "Ancylostoma caninum" or "Ancylostoma braziliense" or "Trichuris tulips" or "Trichuris suis") and Humans)).mp. [mp=title, book title, abstract, original title, name of substance word, subject heading word, floating sub-heading word, keyword heading word, organism supplementary concept word, protocol supplementary concept word, rare disease supplementary concept word, unique identifier, synonyms, population supplementary concept word, anatomy supplementary concept word] | 1254 |
| Web of Science | **((TS=("Ascaris lumbricoides" OR "Trichuris trichiura" OR "Ancyclostoma duodenale" OR "Necator americanus")) AND TS=("small ruminants" OR cow OR cattle OR pig OR swine OR dogs OR canine OR cats OR feline)) OR (TS=("Ascaris suum" OR "Ancylostoma ceylanicum" OR "Ancylostoma caninum" OR "Ancylostoma braziliense" OR "Trichuris vulpis" OR "Trichuris suis") AND TS=Humans)** | 1539 |
| PubMed | (((((("Ascaris lumbricoides") OR ("Trichuris trichiura")) OR ("Ancyclostoma duodenale")) OR ("Necator americanus")) AND ((((("small ruminants") OR (cow OR cattle)) OR (pig OR swine)) OR (dogs OR canine)) OR (cats OR feline))) OR (((((("Ascaris suum") OR ("Ancylostoma ceylanicum")) OR ("Ancylostoma caninum")) OR ("Ancylostoma braziliense")) OR ("Trichuris vulpis")) OR ("Trichuris suis"))) AND (Humans) | 1402 |
