## Supplementary material for "The occurrence of cross-host species soil-transmitted helminth infections in humans and domestic/livestock animals: a systematic review": S2 Table. Reasons for excluding studies..docx

**S2 Table. Reasons for excluding studies after full text assessment against eligibility criteria.**

| **Reasons for exclusion** | **Number of studies** |
| --- | --- |
| Not relevant to our objective | 1011 |
| No PCR | 265 |
| Experimental infection/clinical trial | 53 |
| Did not look for human STH in animals | 42 |
| Did not look for zoonotic STH in animals | 10 |
| Review | 71 |
| Not domestic animal | 69 |
| Not STH species of interest | 32 |
| No full text | 12 |
| Studies with the same sample set as the included studies | 6 |
| **Total number of studies excluded** | **1571** |
