## Supplementary material for "The occurrence of cross-host species soil-transmitted helminth infections in humans and domestic/livestock animals: a systematic review": S3 Table. Description of variables extracted..docx

| **Data extracted** | **Description** |
| --- | --- |
| Authors and Year of publication | Author names, year the study was published |
| Journal title and Article title | Journal title, name of the article |
| Year data were collected | Year when the data collection was done |
| Study design / participants | The design of the study and selection of the participants for the study. For example, cross-sectional study, household selected by systematic sampling and all individuals invited to participate in the study |
| Selection of isolates for molecular testing | The method in which the samples have been selected for molecular testing is noted. For example, all samples collected are subjected to molecular analysis or only microscopy positive samples, or among the microscopy positive samples, some are randomly selected for molecular analysis, etc |
| Numbers tested by morphological examination | The number of samples that were tested by microscopy (whenever applicable) |
| Numbers tested by PCR | The number of samples that were tested by PCR |
| 95% confidence intervals | For survey studies that have provided 95% confidence intervals. |
| PCR method | Molecular method used for species identification. For example, PCR, PCR-RLP, multiplex PCR, etc |
| Gene targets | Gene targets for STH species, for example, ITS-1,2, 5.8s and 28S region, etc |
| Species detected in humans | *Ancylostoma ceylanicum* or *Ancylostoma caninum* or *Ancylostoma braziliense* or *Ascaris suum* or *Trichuris vulpis* or *Trichuris suis* |
| Species detected in animals | *Ascaris lumbricoides* or *Trichuris trichiura* or *Necator americanus* or *Ancylostoma duodenale* |
| Animal hosts | When human STH was detected in animals, the identified animal hosts were noted. |
| Clinical history/symptoms | In the case studies or studies in which the samples have been taken from the patients reported to have symptoms, the associated signs and symptoms are recorded. For example, Abdominal pain, vomiting, constipation, Itchy rash, persistent pruritis |
| Sample collected | Type of sample collected, stool, worms, etc |
| Number of positive cases | From samples that were analysed by PCR , how many were found positive. |
| Travel history/animal contact | In the case studies or studies in which the samples have been taken from the patients reported to have symptoms, any history of travel to an endemic country is recorded. |
