## Supplementary material for "The occurrence of cross-host species soil-transmitted helminth infections in humans and domestic/livestock animals: a systematic review": S4 Table. Quality assessment of studies using the AXIS critical appraisal tool.docx

**S4 Table. Quality assessment of studies using the AXIS critical appraisal tool for cross-sectional studies**[1]**.**

| **Author and year of publication** | 1. **Were the aims/objectives of the study clear?** | 2. **Was the study design appropriate for the stated aims?** | 3. **Was the sample size justified?** | 4. **Was the target/reference population clearly define? Is it clear who the research was about?)** | 5. **Was the sample frame taken from an appropriate population base so that it closely represented the target/reference population** | 6. **Was the selection process likely to select subjects/participants that were representative of the target/reference population under investigation?** | 7. **Were measures undertaken to address and categorise non-responders?** | 8. **Were the risk factor and outcome variables measured appropriate to the aims of the study?** | 9. **Were the risk factor and outcome variables measured correctly using instruments /measurements that had been trialled, piloted or published previously?** | 10. **Is it clear what used to determined statistical significance and /or precision estimates?** | 11. **Were the methods (including statistical methods) sufficiently described to enable them to be repeated?** | 12. **Were the basic data adequately described?** | 13. **Does the response rate raise concern about non -response bias?** | **14. If appropriate, was information about non-response described?** | **15. Were the results internally consistent?** | **16. Were the results presented for all analyses described in the methods?** | **17. Were the authors discussions and conclusion described by the results?** | **18. Were the limitations of the study discussed?** | **19. If the paper describe there were not funding sources or conflicts of interest that may affect the authors' interpretation of the results?** | **20. Was ethical approval or consent of participants attained?** | **Score** |
| --- | --- | --- | --- | --- | --- | --- | --- | --- | --- | --- | --- | --- | --- | --- | --- | --- | --- | --- | --- | --- | --- |
| Aguilar-Rodríguez et al (2024)[2] | 1 | 1 | 1 | 1 | 1 | 1 | 0 | 1 | 1 | 0 | 1 | 1 | 1 | 0 | 1 | 1 | 1 | 1 | 1 | 1 | 17 |
| Agustina et al (2023)[3] | 1 | 1 | 0 | 1 | 1 | 0 | 0 | 1 | 1 | 0 | 1 | 1 | 1 | 0 | 1 | 1 | 1 | 0 | 1 | 1 | 15 |
| Areekul et al (2010)[4] | 1 | 1 | 1 | 1 | 1 | 1 | 0 | 1 | 1 | 1 | 1 | 1 | 1 | 0 | 1 | 1 | 1 | 1 | 1 | 1 | 18 |
| Ash et al (2017)[5] | 1 | 1 | 1 | 1 | 1 | 1 | 0 | 1 | 1 | 1 | 1 | 1 | 1 | 0 | 1 | 1 | 1 | 1 | 1 | 1 | 18 |
| Aula et al (2020)[6] | 1 | 1 | 1 | 1 | 1 | 1 | 0 | 1 | 1 | 1 | 1 | 1 | 1 | 0 | 1 | 1 | 1 | 1 | 1 | 1 | 18 |
| Aung et al (2017)[7] | 1 | 1 | 1 | 1 | 1 | 1 | 0 | 1 | 1 | 1 | 1 | 1 | 1 | 0 | 1 | 1 | 1 | 1 | 1 | 1 | 18 |
| Boyko et al (2020)[8] | 1 | 1 | 1 | 1 | 1 | 1 | 0 | 1 | 1 | 1 | 1 | 1 | 1 | 0 | 1 | 1 | 1 | 1 | 1 | 1 | 18 |
| Bradbury et al (2017)[9] | 1 | 1 | 1 | 1 | 1 | 1 | 0 | 1 | 1 | 1 | 1 | 1 | 1 | 0 | 1 | 1 | 1 | 1 | 1 | 1 | 18 |
| Bui et al (2021)[10] | 1 | 1 | 1 | 1 | 1 | 1 | 0 | 1 | 1 | 1 | 1 | 1 | 1 | 0 | 1 | 1 | 1 | 1 | 1 | 1 | 18 |
| Calvopina et al (2024)[11] | 1 | 1 | 0 | 1 | 1 | 0 | 0 | 1 | 1 | 0 | 1 | 1 | 1 | 0 | 1 | 1 | 1 | 1 | 1 | 1 | 15 |
| Chin et al (2016)[12] | 1 | 1 | 1 | 1 | 1 | 1 | 0 | 1 | 1 | 1 | 1 | 1 | 1 | 0 | 1 | 1 | 1 | 1 | 1 | 1 | 18 |
| Conlan et al (2012)[13] | 1 | 1 | 1 | 1 | 1 | 1 | 0 | 1 | 1 | 1 | 1 | 1 | 1 | 0 | 1 | 1 | 1 | 1 | 1 | 1 | 18 |
| (Colella et al (2021)[14] | 1 | 1 | 1 | 1 | 1 | 1 | 0 | 1 | 1 | 1 | 1 | 1 | 1 | 0 | 1 | 1 | 1 | 1 | 1 | 1 | 18 |
| Dunn et al (2020)[15] | 1 | 1 | 1 | 1 | 1 | 1 | 0 | 1 | 1 | 1 | 1 | 1 | 1 | 0 | 1 | 1 | 1 | 1 | 1 | 1 | 18 |
| George et al (2015)[16] | 1 | 1 | 1 | 1 | 1 | 1 | 0 | 1 | 1 | 1 | 1 | 1 | 1 | 0 | 1 | 1 | 1 | 1 | 1 | 1 | 18 |
| George, Geldhof, et al (2016)[17] | 1 | 1 | 1 | 1 | 1 | 1 | 0 | 1 | 1 | 1 | 1 | 1 | 1 | 0 | 1 | 1 | 1 | 1 | 1 | 1 | 18 |
| George, Levecke, et al (2016)[18] | 1 | 1 | 1 | 1 | 1 | 1 | 0 | 1 | 1 | 1 | 1 | 1 | 1 | 0 | 1 | 1 | 1 | 1 | 1 | 1 | 18 |
| Htun et AL (2021)[19] | 1 | 1 | 1 | 1 | 1 | 1 | 0 | 1 | 1 | 1 | 1 | 1 | 1 | 0 | 1 | 1 | 1 | 1 | 1 | 1 | 18 |
| Hughes et al (2023)[20] | 1 | 1 | 1 | 1 | 1 | 1 | 0 | 1 | 1 | 1 | 1 | 1 | 1 | 0 | 1 | 1 | 1 | 1 | 1 | 1 | 18 |
| Inpankaew et al (2014)[21] | 1 | 1 | 1 | 1 | 1 | 1 | 0 | 1 | 1 | 1 | 1 | 1 | 1 | 0 | 1 | 1 | 1 | 1 | 1 | 1 | 18 |
| Jiraanankul et al (2011)[22] | 1 | 1 | 1 | 1 | 1 | 1 | 0 | 1 | 1 | 1 | 1 | 1 | 1 | 0 | 1 | 1 | 1 | 1 | 1 | 1 | 18 |
| Mohd-Shaharuddin et al (2019)[23] | 1 | 1 | 1 | 1 | 1 | 1 | 0 | 1 | 1 | 1 | 1 | 1 | 1 | 0 | 1 | 1 | 1 | 1 | 1 | 1 | 18 |
| Mulinge et al (2020)  [24] | 1 | 1 | 1 | 1 | 1 | 1 | 0 | 1 | 1 | 1 | 1 | 1 | 1 | 0 | 1 | 1 | 1 | 1 | 1 | 1 | 18 |
| Niamnuy et al (2016)[25] | 1 | 1 | 1 | 1 | 1 | 1 | 0 | 1 | 1 | 1 | 1 | 1 | 1 | 0 | 1 | 1 | 1 | 1 | 1 | 1 | 18 |
| Ngcamphalala et al (2020)[26] | 1 | 1 | 0 | 1 | 1 | 0 | 0 | 1 | 1 | 0 | 1 | 1 | 1 | 0 | 1 | 1 | 1 | 1 | 1 | 1 | 15 |
| Ngui et al (2012)[27] | 1 | 1 | 1 | 1 | 1 | 0 | 0 | 1 | 1 | 1 | 1 | 1 | 1 | 0 | 1 | 1 | 1 | 1 | 1 | 1 | 17 |
| O’Connell et al (2018)[28] | 1 | 1 | 1 | 1 | 1 | 1 | 0 | 1 | 1 | 1 | 1 | 1 | 1 | 0 | 1 | 1 | 1 | 1 | 1 | 1 | 18 |
| Traub et al (2008)[29] | 1 | 1 | 1 | 1 | 1 | 1 | 0 | 1 | 1 | 1 | 1 | 1 | 1 | 0 | 1 | 1 | 1 | 1 | 1 | 1 | 18 |
| Traub et al (2002)[30] | 1 | 1 | 1 | 1 | 1 | 1 | 0 | 1 | 1 | 1 | 1 | 1 | 1 | 0 | 1 | 1 | 1 | 1 | 1 | 1 | 18 |
| Sato et al (2010)[31] | 1 | 1 | 1 | 1 | 1 | 1 | 0 | 1 | 1 | 1 | 1 | 1 | 1 | 0 | 1 | 1 | 1 | 1 | 1 | 1 | 18 |
| Sears et al (2022)[32] | 1 | 1 | 1 | 1 | 1 | 1 | 0 | 1 | 1 | 1 | 1 | 1 | 1 | 0 | 1 | 1 | 1 | 1 | 1 | 1 | 18 |
| Stracke et al (2019)[33] | 1 | 1 | 1 | 1 | 1 | 1 | 0 | 1 | 1 | 1 | 1 | 1 | 1 | 0 | 1 | 1 | 1 | 1 | 1 | 1 | 18 |
| Stracke et al (2021)[34] | 1 | 1 | 1 | 1 | 1 | 1 | 0 | 1 | 1 | 1 | 1 | 1 | 1 | 0 | 1 | 1 | 1 | 1 | 1 | 1 | 18 |
| Webster et al (2022)[35] | 1 | 1 | 1 | 1 | 1 | 1 | 0 | 1 | 1 | 1 | 1 | 1 | 1 | 0 | 1 | 1 | 1 | 1 | 1 | 1 | 18 |

Yes=1, No=0
