## Supplementary material for "The occurrence of cross-host species soil-transmitted helminth infections in humans and domestic/livestock animals: a systematic review": S5 Table. Quality assessment of studies using JBI checklist.docx

**S5 Table. Quality assessment of studies using the JBI checklist for cross-sectional studies**[1]**.**

| **Author and year of publication** | 1. **Were the criteria for inclusion in the sample clearly defined?** | 2. **Were the study subjects and the setting described in detail?** | 3. **Was the exposure measured in a valid and reliable way?** | 4. **Were objective, standard criteria used for measurement of the condition?** | 5. **Were confounding factors identified?** | 6. **Were strategies to deal with confounding factors stated?** | 7. **Were the outcomes measured in a valid and reliable way?** | 8. **Was appropriate statistical analysis used?** | **Score** |
| --- | --- | --- | --- | --- | --- | --- | --- | --- | --- |
| Arizono et al (2010)[2] | 1 | 1 | 1 | 1 | 0 | 0 | 1 | 1 | 6 |
| Furtado et al (2020)[3] | 1 | 1 | 1 | 1 | 0 | 0 | 1 | 1 | 6 |
| Gerber et al (2021)[4] |  |  |  |  |  |  |  |  |  |
| Koehler et al (2013)[5] | 1 | 1 | 1 | 1 | 0 | 0 | 1 | 1 | 6 |
| Phosuk et al (2013)[6] | 1 | 1 | 1 | 1 | 0 | 0 | 1 | 1 | 6 |

Yes=1, No=0
