## Supplementary material for "The occurrence of cross-host species soil-transmitted helminth infections in humans and domestic/livestock animals: a systematic review": S6 Table. Characteristics of eligible studies collected samples from hospitals.docx

**S6 Table: Characteristics of eligible studies that collected samples from hospitals/clinics/laboratories** [1].

| **Study Reference** | **Study location** | **Recruitment method** | **Clinical history/symptoms** | **Method of diagnosis** | **Sample collected** | **Number of samples** | **Species** | **Positive cases** | **Travel history/animal contact** |
| --- | --- | --- | --- | --- | --- | --- | --- | --- | --- |
| Koehler et al (2013)[2] | Australia | Samples of humans with h/o GI disorders were tested in 2 labs | Gastrointestinal disturbances | PCR | Faeces | Amplicons produced in 12/227 | *A.ceylanicum* | 2/12 | - |
| Furtado et al (2020)[3] | Brazilian states | human faeces obtained from commercial laboratories | - | Conventional PCR | Faeces | 634 single hookworm eggs from fecal samples from 53 humans | *A.caninum* | N. americanus, -98·1% (622/634)  Ancylostoma spp.-1·9% (12/634)  Sequencing-  Ancylostoma spp.- 100% similarity with A.caninum, | - |
| Gerber et al (2021)[4] | France | 8 university hospitals (2016-18) Samples of symptomatic patients who have travelled from endemic countries. | Diarrhoea, anemia, hypereosinophilia, abdominal pain | PCR | Faeces | 34 microscopy positive samples from 8 hospitals were analysed by PCR | *A.ceylanicum* | 3/34 | Returned from Pakistan, Cote d‘Ivoire, Colombia, Pakistan and French Guiana |
| Arizono et al (2010)[5] | Japan | Adult ascaris worms passed in faeces/ expelled through mouth or extracted by endoscopy | - | PCR | Adult ascaris worms |  | *A.suum* | *3/9* isolates of pig origin | *-* |
| Phosuk et al (2013)[6] | Thailand | Samples from 10 patients from a hospital and 20 from villagers southern Thailand | - | PCR | Faeces | 30 | *A.ceylanicum* | 3/30 | - |
