## Supplementary material for "The occurrence of cross-host species soil-transmitted helminth infections in humans and domestic/livestock animals: a systematic review": S7 Table. Quality assessment of case reports.docx

### **S7 Table. Quality assessment of case reports using the JBI checklist for case reports.**

| **Author and year of publication** | 1. **Were patient’s demographic characteristics clearly described?** | 2. **Was the patient’s history clearly described and presented as a timeline?** | 3. **Was the current clinical condition of the patient on presentation clearly described?** | 4. **Were diagnostic tests or assessment methods and the results clearly described?** | 5. **Was the intervention(s) or treatment procedure(s) clearly described?** | 6. **Was the post-intervention clinical condition clearly described?** | 7. **Were adverse events (harms) or unanticipated events identified and described?** | 8. **Does the case report provide takeaway lessons?** | **Score** |
| --- | --- | --- | --- | --- | --- | --- | --- | --- | --- |
| Brunet et al (2015)[1] | 1 | 1 | 1 | 1 | 1 | 1 | 0 | 1 | 7 |
| Dutto & Petrosillo (2013)[2] | 1 | 1 | 1 | 1 | 1 | 1 | 0 | 1 | 7 |
| Nath et al (2024)[3] | 1 | 1 | 1 | 1 | 0 | 0 | 0 | 1 | 5 |
| Nishioka et al (2024)[4] | 1 | 1 | 1 | 1 | 1 | 1 | 0 | 1 | 7 |
| Poppert et al (2017)[5] | 1 | 1 | 1 | 1 | 1 | 1 | 1 | 1 | 8 |
| Le Joncour et al (2012)[6] | 1 | 1 | 1 | 1 | 1 | 1 | 0 | 1 | 7 |
| Romano et al (2021)[7] | 1 | 1 | 1 | 1 | 1 | 1 | 0 | 1 | 7 |
| Jung et al (2020)[8] | 1 | 1 | 1 | 1 | 1 | 0 | 0 | 1 | 6 |
| Kaya et al (2016)[9] | 1 | 1 | 1 | 1 | 1 | 1 | 0 | 1 | 7 |
| Ngui et al (2014)[10] | 1 | 1 | 1 | 1 | 0 | 0 | 0 | 1 | 5 |
| Yoshikawa et al (2018)[11] | 1 | 1 | 1 | 1 | 1 | 1 | 0 | 1 | 7 |

Yes=1, No=0
