## Supplementary figures and images for "The occurrence of cross-host species soil-transmitted helminth infections in humans and domestic/livestock animals: a systematic review"

### S1 Fig. Number of studies per year and the cumulative total of included studies.docx

**S1 Fig. Number of studies per year and the cumulative total of included studies.**
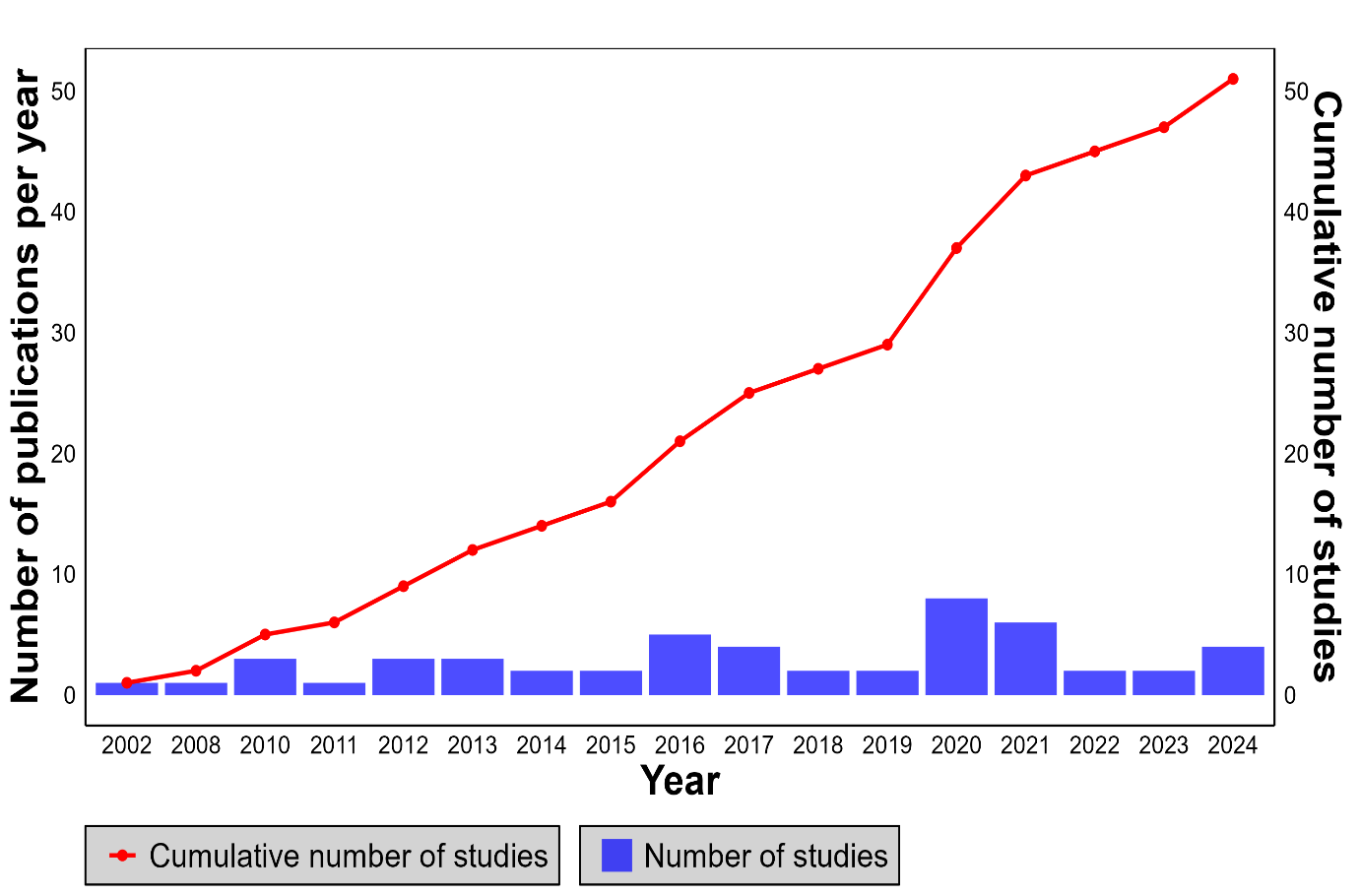

### S2 Fig. Number of studies by country reporting zoonotic STH in humans..docx

**S2 Fig. Number of studies by country reporting zoonotic STH in humans.**

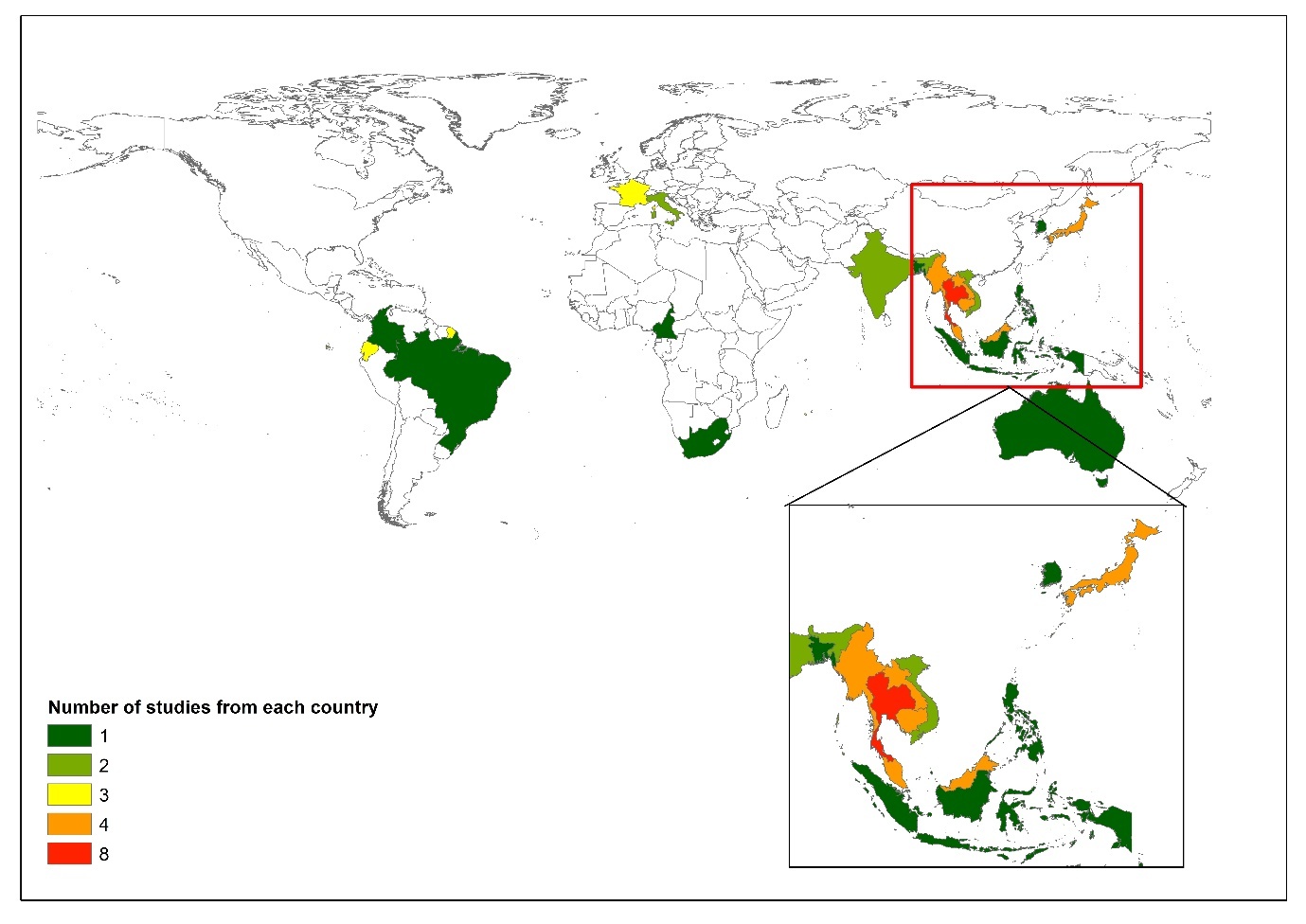

### S3 Fig. Number of studies by country reporting human STH in animals..docx

**S3 Fig: Number of studies by country reporting human STH in animals.**

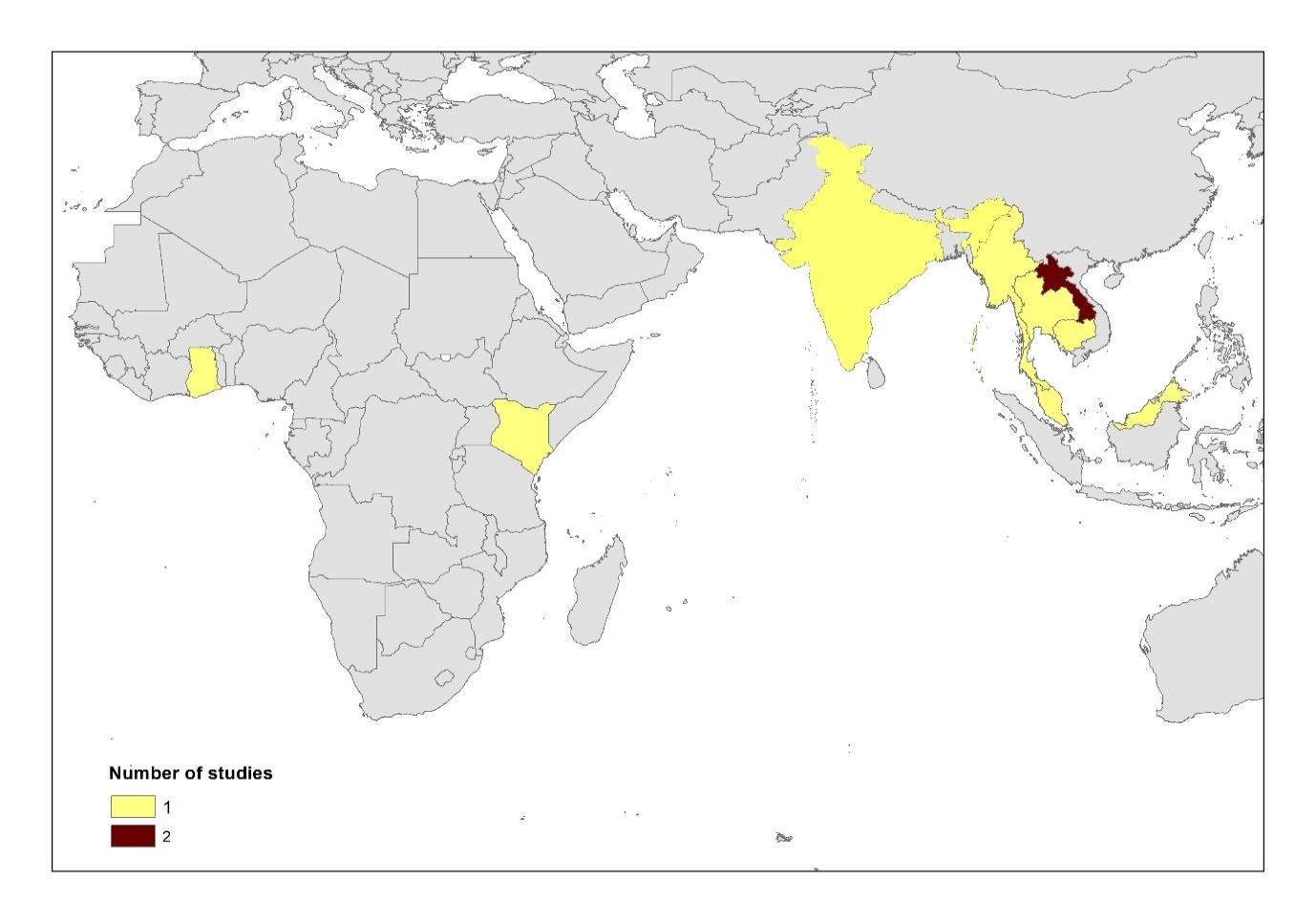
